# Graft-versus-host disease prophylaxis shapes T cell biology and immune reconstitution after hematopoietic cell transplant

**DOI:** 10.1101/2025.02.25.25322901

**Authors:** Steven J. Siegel, Susan DeWolf, Joseph Schmalz, Wael Saber, Jiayi Dong, Michael J. Martens, Brent Logan, Alexandre Albanese, Lorenzo Iovino, Edward Chen, James Kaminski, Donna Neuberg, Kyle Hebert, Paula Keskula, Jillian Zavistaski, Lea Steinberg, Isabella Schichter, Lorenzo Cagnin, Vanessa Hernandez, Makya Warren, Kristy Applegate, Merav Bar, Saurabh Chhabra, Sung Won Choi, William Clark, Suman Das, Robert Jenq, Richard J. Jones, John E. Levine, Hemant Murthy, Armin Rashidi, Marcie Riches, Karamjeet Sandhu, Anthony D. Sung, Karilyn Larkin, Monzr M. Al Malki, Mahasweta Gooptu, Hany Elmariah, Amin Alousi, Lyndsey Runaas, Brian Shaffer, Andrew Rezvani, Najla El Jurdi, Alison W. Loren, Danielle Scheffey, Catherine Sanders, Mehdi Hamadani, Jarrod Dudakov, Stephanie Bien, Harlan Robins, Mary Horowitz, Javier Bolaños-Meade, Shernan Holtan, Ami S. Bhatt, Miguel-Angel Perales, Leslie S. Kean

**Affiliations:** Department of Pediatrics, Harvard Medical School and Divisions of Infectious Diseases and Hematology/Oncology, Boston Children’s Hospital, Boston, MA; Leukemia Service, Memorial Sloan Kettering Cancer Center, New York, NY; Adaptive Biotechnologies, Seattle, WA; Department of Medicine, Medical College of Wisconsin and Center for International Blood and Marrow Transplant Research (CIBMTR), Milwaukee, WI; Division of Hematology/Oncology, Boston Children’s Hospital, Boston, MA; CIBMTR and Institute for Health and Equity, Medical College of Wisconsin, Milwaukee, WI; CIBMTR and Division of Biostatistics, Medical College of Wisconsin, Milwaukee, WI; Clinical Research Division, Fred Hutchinson Cancer Center, Seattle, WA; Division of Pediatric Hematology/Oncology, Boston Children’s Hospital, Boston, MA and the Broad Institute of the Massachusetts Institute of Technology and Harvard, Cambridge, MA; Department of Data Science, Dana-Farber Cancer Institute, Boston, MA; Translational Science and Therapeutics Division, Fred Hutchinson Cancer Center, Seattle, WA; Emmes, Rockville, MD; Division of Hematology/Oncology, Mayo Clinic, Phoenix, AZ; Department of Pediatrics, University of Michigan, Ann Arbor, MI; Division of Hematology–Oncology and Palliative Care, Virginia Commonwealth University, Richmond, VA; Department of Medicine, Vanderbilt University School of Medicine, Nashville, TN; Department of Hematology and Hematopoietic Cell Transplantation, City of Hope, Duarte, CA; Sidney Kimmel Comprehensive Cancer Center at Johns Hopkins University and the Department of Oncology, Johns Hopkins University School of Medicine, Baltimore, MD; Tisch Cancer Institute, Icahn School of Medicine at Mount Sinai, New York, NY; Division of Hematology-Oncology and Blood and Marrow Transplant and Cellular Therapy Programs, Mayo Clinic, Jacksonville, FL; CIBMTR, Milwaukee, WI; Division of Hematologic Malignancies and Cellular Therapy, Kansas University Medical Center, Kansas City, KS; Ohio State University Comprehensive Cancer Center, Columbus, OH; Department of Hematology and Oncology, Dana-Farber Cancer Institute, Boston, MA; Department of Blood and Marrow Transplant and Cellular Immunotherapy, H. Lee Moffitt Cancer and Research Institute, Tampa, FL; Department of Stem Cell Transplantation and Cellular Therapy, the University of Texas M.D. Anderson Cancer Center, Houston, TX; Division of Hematology and Oncology, Medical College of Wisconsin, Milwaukee, WI; Adult Bone Marrow Transplantation Service, Memorial Sloan Kettering Cancer Center, and the Department of Medicine, Weill Cornell Medical College, New York, NY; Division of Blood and Marrow Transplantation and Cellular Therapy, Stanford University School of Medicine, Stanford, CA; Center for Cancer Research, National Cancer Institute, Bethesda, MD; Division of Hematology and Oncology, Perelman School of Medicine, University of Pennsylvania, Philadelphia, PA; Blood and Marrow Transplant Program and Cellular Therapy Program, Medical College of Wisconsin and CIBMTR, Milwaukee, WI; Translational Science and Therapeutics Division, Fred Hutchinson Cancer Center and Department of Immunology, University of Washington, Seattle, WA; Blood and Marrow Transplantation Section, Roswell Park Comprehensive Cancer Center, Buffalo, NY; Division of Hematology, Departments of Medicine and Genetics, Stanford University, Palo Alto, CA; Department of Pediatrics, Harvard Medical School, Division of Hematology/Oncology, Boston Children’s Hospital, and Department of Pediatric Oncology, Dana-Farber Cancer Institute, Boston, MA

## Abstract

Successful hematopoietic cell transplant requires immunosuppression to prevent graft-versus-host disease (GVHD), a lethal, T-cell-mediated post-transplant complication. The phase 3 BMT CTN 1703 trial demonstrated superior GVHD-free/relapse-free survival for post-transplant cyclophosphamide (PT-Cy)-based GVHD prophylaxis versus tacrolimus/methotrexate (Tac/MTX), but did not improve overall survival. To compare T-cell biology between GVHD prophylaxis regimens, 324 patients were co-enrolled onto BMT CTN 1801 (NCT03959241). We quantified T-cell immune reconstitution using multi-modal analysis, including T-cell receptor (TCR) sequencing of 2,359 longitudinal samples (180,432,350 T-cells). Compared to Tac/MTX, PT-Cy was associated with an early, substantial reduction in TCR diversity that was sustained for 2 years. PT-Cy led to a T-cell reconstitution bottleneck, including reduced thymic output and virus-associated TCRs. Decreased D+14 TCR diversity predicted prevention of chronic GVHD, but also correlated with increased moderate-to-severe infections. This study reveals how distinct immunosuppression strategies have significant effects on the global immune repertoire, underpinning post-transplant clinical outcomes.

## Introduction

Immunosuppression to prevent graft-versus-host disease (GVHD) is fundamental to the efficacy of allogeneic hematopoietic cell transplantation (HCT).^1–5^ However, T cell-directed immunomodulation creates a clinical paradox: while control of T-cell expansion and activation is essential to mitigate GVHD, appropriate reconstitution of the circulating T-cell pool is critical for recovery of robust immunity post-transplant.^6^

Several immunosuppressive strategies exist for GVHD prophylaxis.^2,7–12^ The combination of tacrolimus and methotrexate (Tac/MTX) was historically the most common approach.^13,14^ Post-transplant cyclophosphamide (PT-Cy)-based prophylaxis, as tested in a previously-reported Phase 3 trial (BMT CTN 1703),^15^ recently emerged as a new standard for reduced-intensity unrelated-donor HCT, predominantly due to its salutary effects on acute and chronic GVHD (cGVHD).

Results of the BMT CTN 1703 trial demonstrated that PT-Cy significantly improved GVHD-associated outcomes at 1 year post-HCT compared to Tac/MTX.^15^ One-year overall survival, non-relapse mortality and disease-free survival did not differ significantly between the study arms, however, and grade 2 (moderate) infections defined by BMT CTN grading^16^ were more frequent with PT-Cy. The two-year overall survival and disease-free survival also did not differ significantly (Holtan SG, Bolaños-Meade J, Al Malki MM, et al. Improved Patient-Reported Outcomes with Post-Transplant Cyclophosphamide: A Quality-of-Life Evaluation of BMT CTN 1703. Journal of Clinical Oncology *In Press*). To investigate the biologic underpinnings of these results, 324 patients were co-enrolled onto BMT CTN 1801, a landmark biology study. Here, we present findings of this study, which identify T-cell diversity as a key underlying biological determinant of the immunologic effects of PT-Cy, and of transplant outcomes. They introduce a paradigm shift in our understanding of the key role that early T-cell diversity plays in shaping the long-term outcomes of HCT.

## Results

### Patient characteristics

A total of 324 patients enrolled on BMT CTN 1703^15^ were co-enrolled onto this study (165 from the PT-Cy arm and 159 from the Tac/MTX arm). To quantify T-cell diversity post-HCT, we measured the TCR repertoire using high-throughput deep sequencing of TCR beta-chains (timepoints listed in **Supplementary Table 1,** samples obtained shown in **Extended Data Figure 1**). This methodology identifies the scope of TCR beta chains expressed by T cells, a key metric for T-cell immune competence^17,18^ (**Figure 1**). A subset of patients also underwent analysis with multiparameter flow cytometry (**Supplementary Table 2**; n=40, 20 per treatment arm) at the same time-points used for TCR-Seq, with 13 of these patients also undergoing linked single-cell RNA-and TCR Sequencing (on Days 28 and 365 post-HCT, **Supplementary Table 2**).

**Figure 1:**
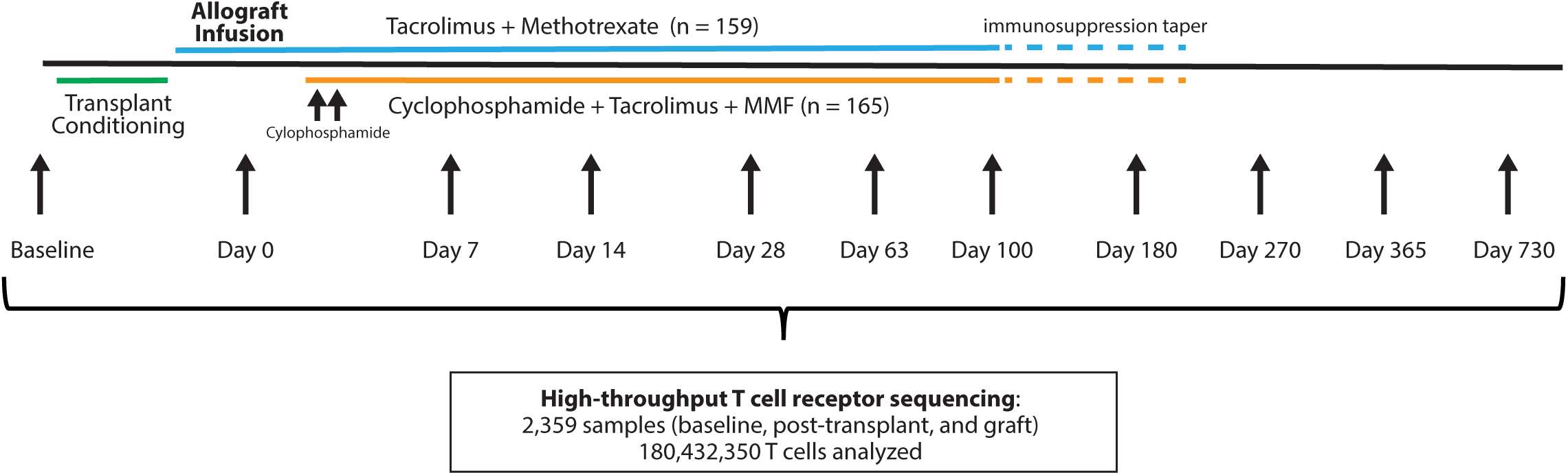
Overview of the BMT CTN 1801 study. Patients were co-enrolled with the BMT CTN 1703 study that randomized GVHD prophylaxis to Tac/MTX or PT-Cy. Peripheral blood samples were obtained longitudinally and TCR-sequencing used to assess the diversity of T cell receptors present after transplant.

All patients received reduced-intensity conditioning prior to HCT for hematologic malignancies. Transplant characteristics and outcomes were representative of the 1703 companion trial (**Table 1** and **Extended Data Table 1**). All patients received peripheral blood stem cell transplants, the majority (67%) from HLA-matched unrelated donors. The primary endpoint for the BMT CTN 1703 trial, GRFS, was higher in patients receiving PT-Cy. Overall survival and the cumulative incidence of disease relapse were not significantly different between the study arms. Grade 2 (moderate) infections^16^ were more frequent in PT-Cy recipients, whereas the cumulative incidences of cGVHD requiring immunosuppression, and severe acute (grade III to IV) GVHD were higher with Tac/MTX, as detailed previously.^15^

**Table 1.** Characteristics and clinical outcomes of patients.

| **Table 1. Characteristics and clinical outcomes of patients** |  |  |  |
| --- | --- | --- | --- |
| **Characteristic** | **Tac/MTX group**<br>**(N=159)** | **PT-Cy group**<br>**(N=165)** | **All patients**<br>**(N=324)** |
| Age – mean in yrs ± SD | 64.6 ± 8.2 | 64.4 ± 8.2 | 64.5 ± 8.2 |
| Male sex – no. (%) | 91 (57.2) | 99 (60.0) | 190 (58.6) |
| Race or ethnic group – no. (%) |  |  |  |
| Hispanic or Latinx ethnic group |  |  |  |
| Hispanic or Latinx | 15 (9.4) | 7 (4.2) | 22 (6.8) |
| Not Hispanic or Latinx | 143 (89.9) | 156 (94.5) | 299 (92.3) |
| Not reported or unknown | 1 (0.6) | 2 (1.2) | 3 (0.9) |
| American Indian or Alaska Native | 1 (0.6) | 0 | 1 (0.3) |
| Asian | 3 (1.9) | 8 (4.8) | 11 (3.4) |
| Black | 4 (2.5) | 8 (4.8) | 12 (3.7) |
| Native Hawaiian or Pacific Islander | 0 | 0 | 0 |
| White | 142 (89.3) | 139 (84.2) | 281 (86.7) |
| Multiple | 0 | 0 | 0 |
| Unknown | 9 (5.7) | 10 (6.1) | 19 (5.9) |
| Primary disease – no. (%) |  |  |  |
| Acute lymphoblastic leukemia | 18 (11.3) | 9 (5.5) | 27 (8.3) |
| Acute myeloid leukemia | 74 (46.5) | 83 (50.3) | 157 (48.5) |
| Myelodysplastic syndrome | 47 (29.6) | 53 (32.1) | 100 (30.9) |
| Other | 20 (12.6) | 20 (12.1) | 40 (12.3) |
| Donor type and HLA matching – no. (%) |  |  |  |
| Related donor 6/6 | 49 (30.8) | 46 (27.9) | 95 (29.3) |
| Unrelated donor 8/8 | 104 (65.4) | 114 (69.1) | 218 (67.3) |
| Unrelated donor 7/8 | 6 (3.8) | 5 (3.0) | 11 (3.4) |
| CMV status (Donor/recipient) – no. (%) |  |  |  |
| -/- | 52 (32.7) | 43 (26.1) | 95 (29.3) |
| +/- | 24 (15.1) | 15 (9.1) | 39 (12.0) |
| -/+ | 28 (17.6) | 54 (32.7) | 82 (25.3) |
| +/+ | 52 (32.7) | 49 (29.7) | 101 (31.2) |
| Missing/unknown | 3 (1.9) | 4 (2.4) | 7 (2.2) |
| **Outcome** | **Percent (95% confidence interval)** |  |  |
| GVHD-free, relapse-free survival | 39.0 (31.3—46.7) | 58.8 (51.2—66.4) | 49.1 (43.6—54.6) |
| Cumulative incidence of acute GVHD |  |  |  |
| Grade II to IV | 54.7 (46.9—62.5) | 50.9 (43.2—58.6) | 52.3 (47.3—58.2) |
| Grade III or IV | 17.0 (11.1—22.9) | 6.1 (2.4—9.7) | 11.4 (7.9—14.9) |
| Cumulative incidence of chronic GVHD at 1 yr | 33.3 (25.9—40.7) | 18.2 (12.2—24.1) | 25.6 (20.8—30.4) |
| Cumulative incidence of infection at 1 yr |  |  |  |
| Grade 2 or 3 | 32.1 (24.7—39.4) | 37.0 (29.5—44.4) | 34.6 (29.4—39.8) |
| Grade 2 | 18.2 (12.2—24.3) | 26.1 (19.3—32.8) | 22.2 (17.7—26.8) |
| Grade 3 | 10.7 (5.8—15.6) | 9.1 (4.7—13.5) | 9.9 (6.6—13.1) |
| Cumulative incidence of graft failure at 1 yr |  |  |  |
| Primary | 1.9 (0—4.0) | 2.4 (0.1—4.8) | 2.2 (0.6—3.8) |
| Secondary | 0.6 (0—1.9) | 3.6 (0.8—6.5) | 2.2 (0.6—3.8) |
| Cumulative incidence of 1-yr relapse and progression | 21.4 (14.9—27.8) | 17.6 (11.7—23.4) | 19.4 (15.1—23.8) |
| Overall survival at 1 yr | 74.8 (68.1—81.7) | 81.2 (75.2—87.2) | 78.1 (73.6—82.6) |

### Association between T cell repertoire diversity and post-transplant outcomes

To compare the composition and diversity of the TCR repertoire – the total population of T-cell clones circulating in an individual defined by their unique set of TCRs – we performed high-throughput TCR sequencing of 2,359 samples and 180,432,350 T cells, with a median of 44,077 profiled TCRs/sample. While T cell diversity within the grafts themselves were not different between groups (**Figure 2a**), after transplant, PT-Cy-treated patients demonstrated significantly reduced TCR repertoire size and diversity versus Tac/MTX. These differences were evident as early as 7 days post-transplant and persisted for up to 2 years across multiple quantitative diversity measures,^19^ including inverse Simpson diversity, Shannon entropy, richness, and singleton TCRs (“singletons” compare single T-cell clones downsampled to adjust for the number of TCRs sequenced, which was robust to a full range of downsampling thresholds (**Figure 2b** and **Extended Data Figures 2-3**). These results demonstrate that the divergence in TCR diversity in Tac/MTX versus PT-Cy was substantial, sustained, and evident early after transplant (with the greatest differences between arms at Day +14, **Supplementary Table 3)**, which raised the hypothesis that early measures of TCR diversity may predict transplant outcomes.

**Figure 2:**
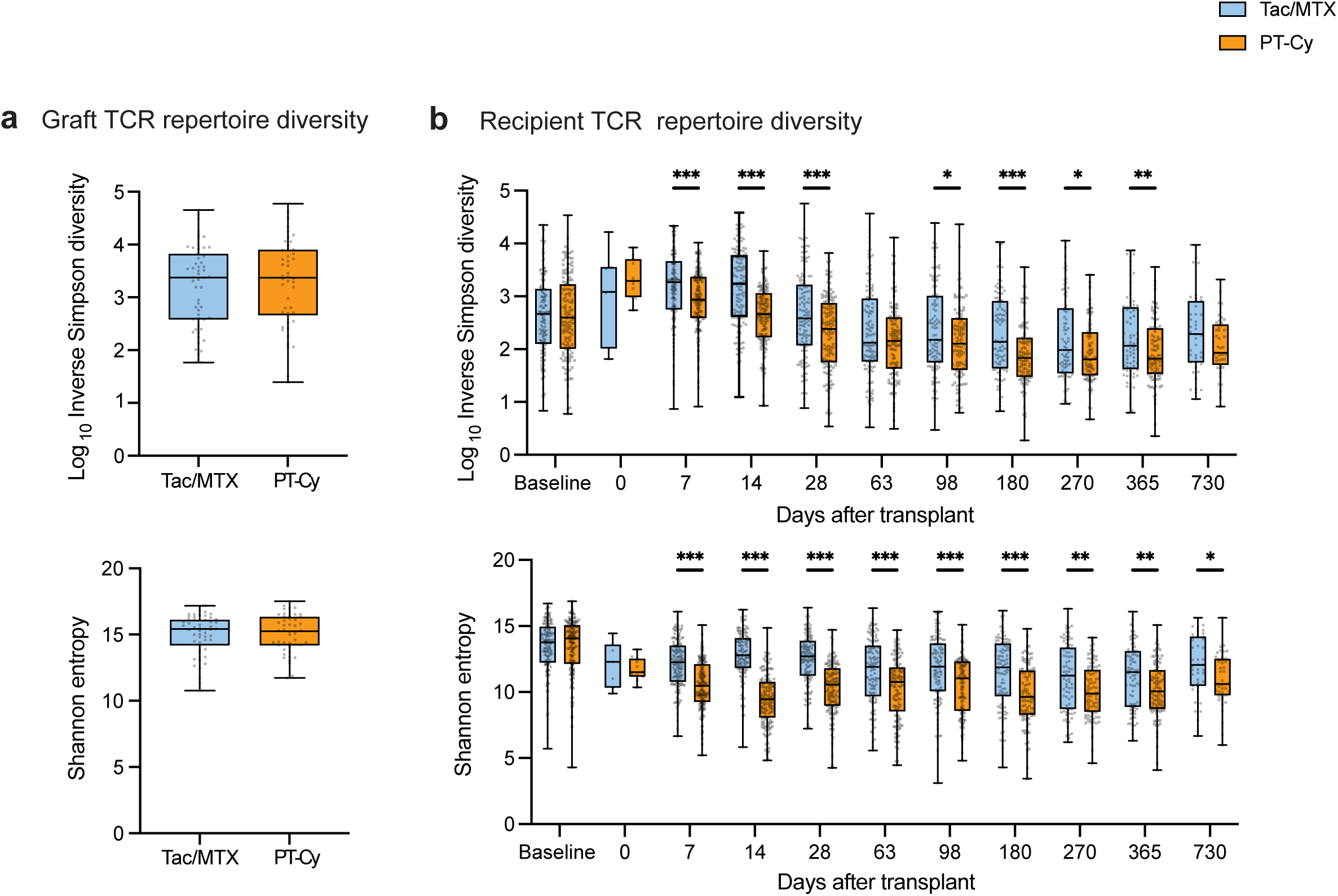
PT-Cy impairs the size and diversity of the T-cell repertoire. a, Global TCR repertoire diversity as measured by inverse Simpson diversity and Shannon entropy was calculated for graft samples (N=101). Comparisons between treatment arms were made using Mann-Whitney U tests. **b,** Global TCR repertoire diversity as measured by inverse Simpson diversity (left) and Shannon entropy (right) was calculated for each patient at each timepoint (N=324 patients). Log-normalized values were compared between arms using a mixed effects model for repeated measures testing, followed by Fisher’s least significant difference posttests at each timepoint. Asterisks indicate P values for these comparisons, * P<0.05, ** P<0.01, *** P<0.001.

To test this possibility, we stratified all patients – independent of treatment arm – into TCR diversity categories and tested whether TCR diversity itself could predict several of the key transplant outcomes measured in the 1703 study. These results demonstrated that while later time-points (Day 98 and 180) were not predictive of cGVHD and infections (**Supplementary Tables 4-5**), early TCR diversity (measured at Day 14 post-HCT, the timepoint of greatest divergence in the Tac/MTX versus PT-Cy arms) was associated with both outcomes: **Figure 3a** demonstrates that patients with low (below the median) versus high (above the median) diversity (**Supplementary Table 6)** had a lower cumulative incidence of cGVHD requiring immunosuppression (28.9% for higher diversity patients vs. 18.2% for lower diversity patients, 95% confidence intervals [CIs] 21.5 to 36.7 and 12.2 to 25.1, respectively; P=0.035). Early TCR diversity was also associated with the cumulative incidence of grade 2 to 3 (moderate to severe/life-threatening) infections, with higher TCR diversity protective against infection (27.5% for higher diversity patients vs. 45.0% for lower diversity patients, 95% CIs 20.3 to 35.1 and 36.6 to 53.1, respectively; P=0.004)(**Figure 3b** and **Extended Data Table 2**), and higher TCR diversity also leading to improved infection-free survival (**Figure 3c).** Importantly, the incidence of chronic GVHD and Grade 2-3 infections followed similar diversity patterns within each treatment arm, though this analysis had limited power when separated by arm (**Extended Data Figure 4a-b**). Lower NRM was observed for patients with higher TCR diversity, but the difference did not reach statistical significance (P=0.05, **Extended Data Figure 4c**). Day 14 inverse Simpson diversity did not predict overall survival (P=0.34, **Figure 3d**), consistent with lower TCR diversity being linked to both positive (protection against cGVHD) and negative (infection risk) transplant outcomes. It also did not predict relapse risk, (P=0.16, **Extended Data Figure 4d**) consistent with similar relapse rates measured in the BMT CTN 1703 trial for patients treated with Tac/MTX versus PT-Cy.^15^

**Figure 3:**
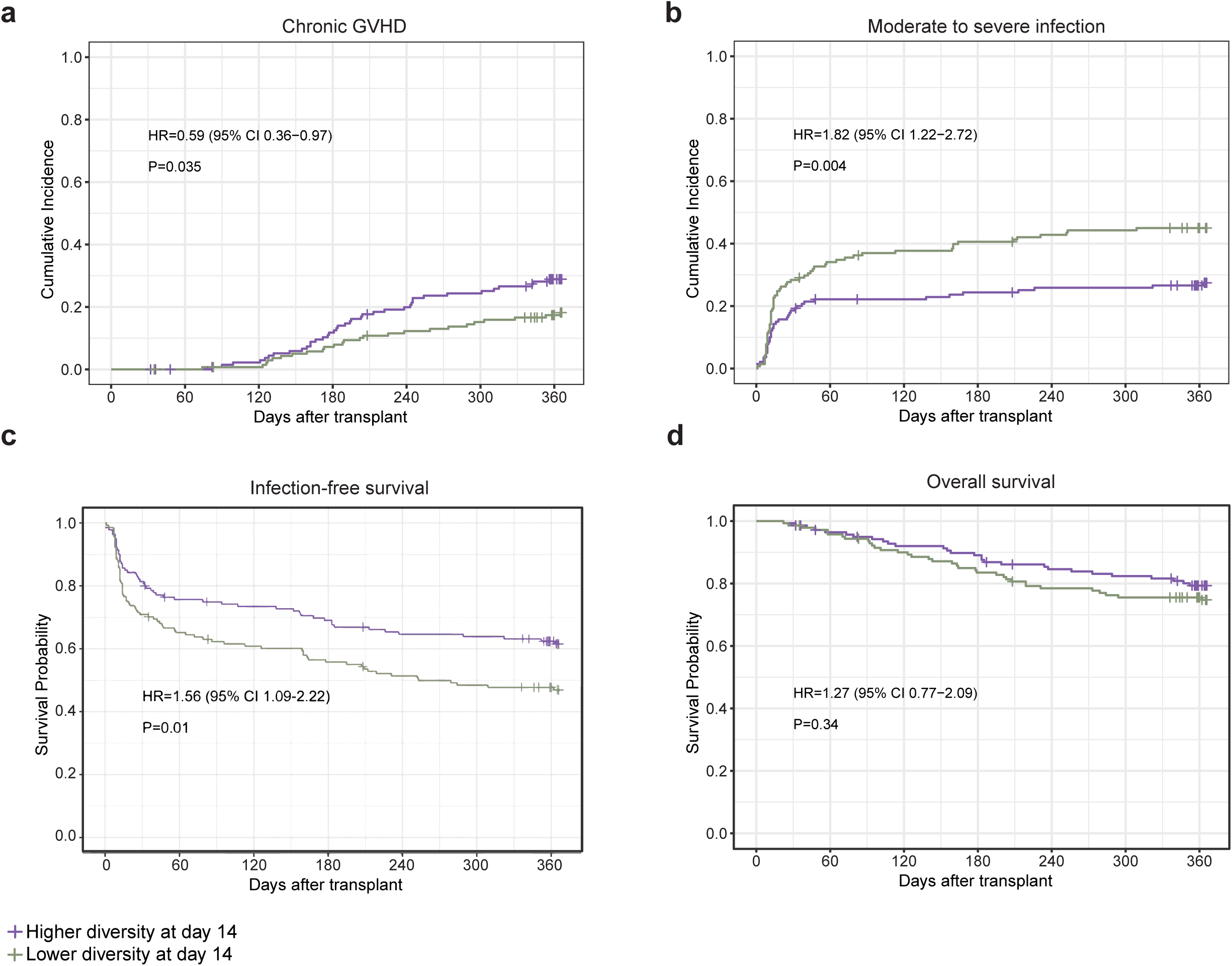
TCR diversity correlates with post-transplant clinical outcomes. In **a** (chronic GVHD requiring systemic immunosuppression) and **b** (moderate to severe infection), all patients with Day 14 samples available (N=281 patients) were dichotomized into high and low diversity groups using the median inverse Simpson diversity index value. The cumulative incidence of chronic GVHD and infection were plotted through 1 year after transplant. The 1-year estimates of the cumulative incidences are provided by group along with 95% confidence intervals. Gray’s test was used for comparisons between high and low diversity groups. Univariate Fine-Gray regression models were also fit for each outcome and set of diversity groups and used to present a subdistribution hazard ratio for the lower diversity group relative to the higher diversity group. The competing risk for chronic GVHD and infection was death in the absence of the applicable outcome. **c,** Survival without moderate or severe infection was also plotted through 1 year after transplant using Kaplan-Meier curves. Log-rank test was used for group comparisons and a univariate Cox regression model fit to present a subdistribution hazard ratio. **d,** Overall survival was assessed for patients dichotomized into high and low diversity groups (N=281 patients) based on median day 14 inverse Simpson diversity and compared using the Kaplan-Meier estimator with a log rank test. All endpoints were censored at 1-year post-transplant or last follow-up, whichever occurred first.

### T-cell repertoire diversity bottleneck emerges following PT-Cy

We next determined which T-cell subsets were altered after PT-Cy. We found a substantial reduction in the size of the T-cell population, with pan-T cell depletion measured by flow cytometry (mean number of T cells/μL at 14 days post-transplant 239.0 for Tac/MTX vs. 19.5 for PT-Cy, P=0.0054, **Figure 4a**). PT-Cy resulted in a significant reduction in all T-cell populations (including total T cells, CD4+ and CD8+ T cells and their subpopulations, CD4 Tregs and γδ T cells) (**Figure 4b-d, Extended Data Figure 5-6**). This reduction was specific to T cells and not observed for NK or B cells (**Extended Data Figure 5**). This reduction was most notable early, before Day +60. While all T-cell populations were depleted early after PT-Cy, this regimen resulted in a particularly prolonged deficit in naive CD4 and CD8 T cells compared to Tac/MTX, which persisted for 2 years post-HCT (**Figure 4b-c**; P=0.002 for CD4 naïve T cells and P<0.001 for CD8 naive T cells).

**Figure 4:**
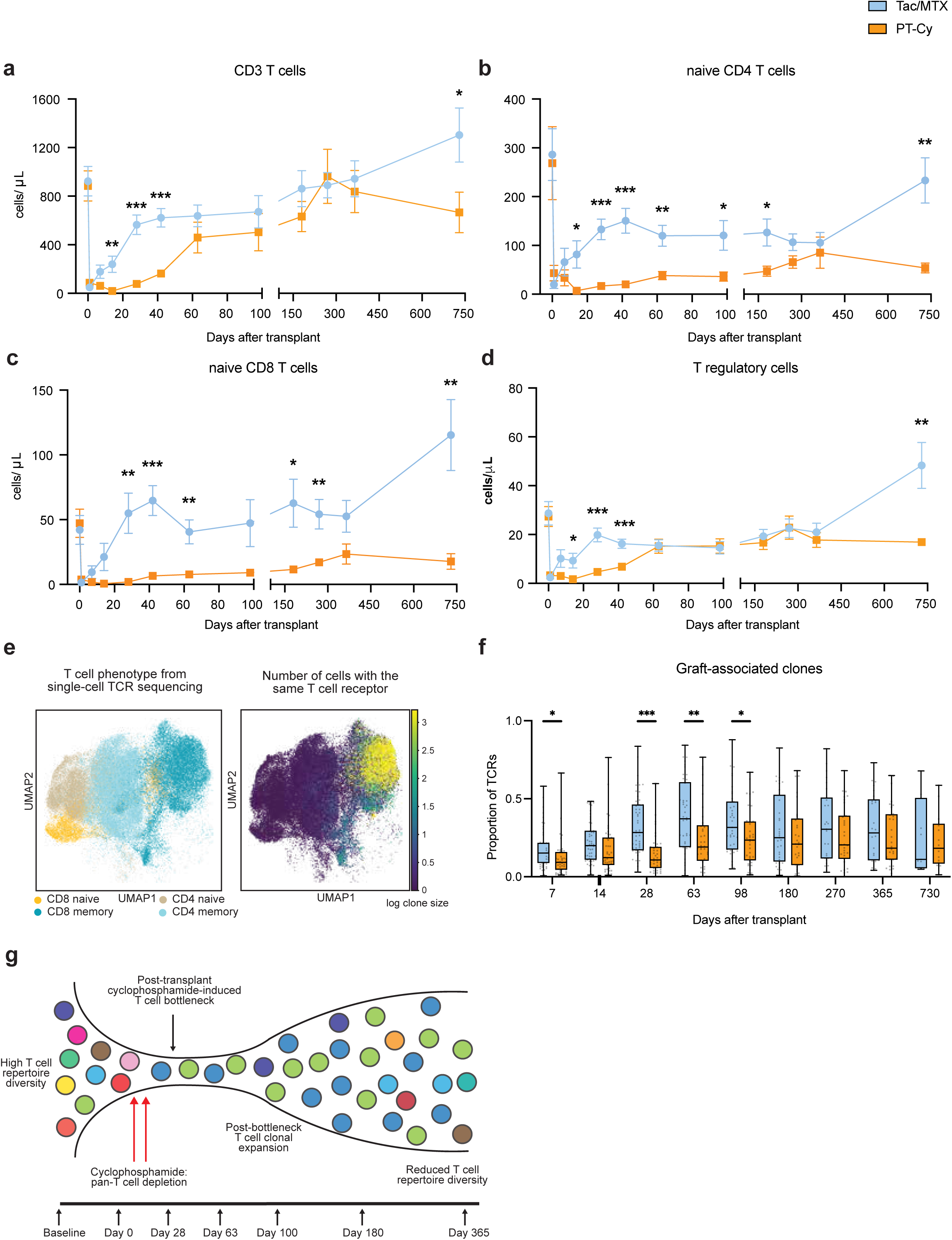
GVHD prophylaxis shapes the composition of immune reconstitution post-transplant. The number of T cells (**a**), naive CD4 (**b**) and CD8 T cells (**c**) and CD4 regulatory T cells (**d**) were measured by flow cytometry (N=40 patients). Data are presented as mean±SEM, and values were compared between arms at each timepoint using mixed effects models for repeated measures testing with Fisher’s least significant difference posttests at each timepoint. Asterisks indicate P values for these comparisons, * P<0.05, ** P<0.01, *** P<0.001. **e,** UMAP visualization of single cell RNA sequencing of day 28 and day 365 (N=8 for PT-Cy and N=5 for Tac/MTX). T cells colored by T cell subsets (CD4 vs CD8, naïve vs memory) (left). UMAP of T cells colored by the size of the TCR clone detected (log scale) (right). TCR clones were identified as being from the stem cell graft based on annotation of sequences from the leftover graft bag/syringe. **f,** The frequency of graft-derived clones at the indicated timepoints was calculated for each patient who had a graft sample available for comparison (N=101 patients). **g,** Schematic of the T cell bottleneck effect following treatment with PT-Cy.

We next performed combined single-cell RNA/TCR-sequencing to link the phenotype of individual T cells to specific TCR clonotypes. We sampled T-cell clones at day 28 (n = 13) and day 365 (n = 13) post-transplant. After identifying naïve and memory T-cell subsets transcriptionally (**Figure 4e** and **Extended Data Figure 7a**), we evaluated the distribution of TCRs across phenotypic compartments. Expanded TCRs, those clonotypes present in larger numbers in a given sample, were found exclusively in memory T-cell subsets. Naïve cell subsets, by contrast, were mostly singleton TCRs (**Figure 4e**). We were able to identify clones captured in both single-cell and bulk TCR sequencing analysis, with similar results (**Extended Data Figure 7b)**.

Of note, for both Tac/MTX and PT-Cy patients, a significant proportion of T cells from the donor graft contributed to the T-cell compartment for years after transplant (**Figure 4f)**.

However, this proportion was significantly higher in Tac/MTX compared to PT-Cy early post-transplant, consistent with early depletion of graft-associated clones arising from cyclophosphamide treatment.

Taken together, these findings suggest that HCT results in a bottleneck effect^20–23^ on the T-cell repertoire, particularly in PT-Cy relative to Tac/MTX (**Figure 4g**). A bottleneck occurs when there is a sharp reduction in the size of a population due to external events (such as post-transplant treatment with cyclophosphamide) that results in a significant reduction in that population’s genetic variation^20–22^, here manifesting as reduced TCR diversity. This bottleneck affected all T-cell subsets along with graft-associated TCRs, and likely is a critical component of the mechanism underpinning the reduction in alloreactive T cells that mediate GVHD with PT-Cy.

### Release of the TCR repertoire bottleneck

After suppression of the T-cell population by PT-Cy, there was delayed expansion of most of the remaining T-cell populations. This was measured by several complementary approaches. First, T-cell proliferation (Ki67 expression) was measured using flow cytometry, which documented a delayed pattern of T-cell proliferation with PT-Cy versus Tac/MTX (for example, naïve CD4 T-cell proliferation peaked at Day 7 with Tac/MTX versus Day 42 with PT-Cy, difference between arms P=0.01 at Day 7 and P=0.008 at Day 42, **Figure 5a**). The delayed T-cell expansion after PT-Cy was associated with delayed TCR repertoire turnover, detected by quantifying the sharing of TCR clones between successive timepoints using the Morisita Index, a normalized metric ranging from 0 to 1 (with 1 denoting complete repertoire overlap). This analysis revealed that Tac/MTX-treated patients reached a stable TCR repertoire more quickly than PT-Cy treated patients (**Figure 5b**), with the Morisita > 0.5 by Day 14 (Tac/MTX) versus Day 63 (PT-Cy).

**Figure 5:**
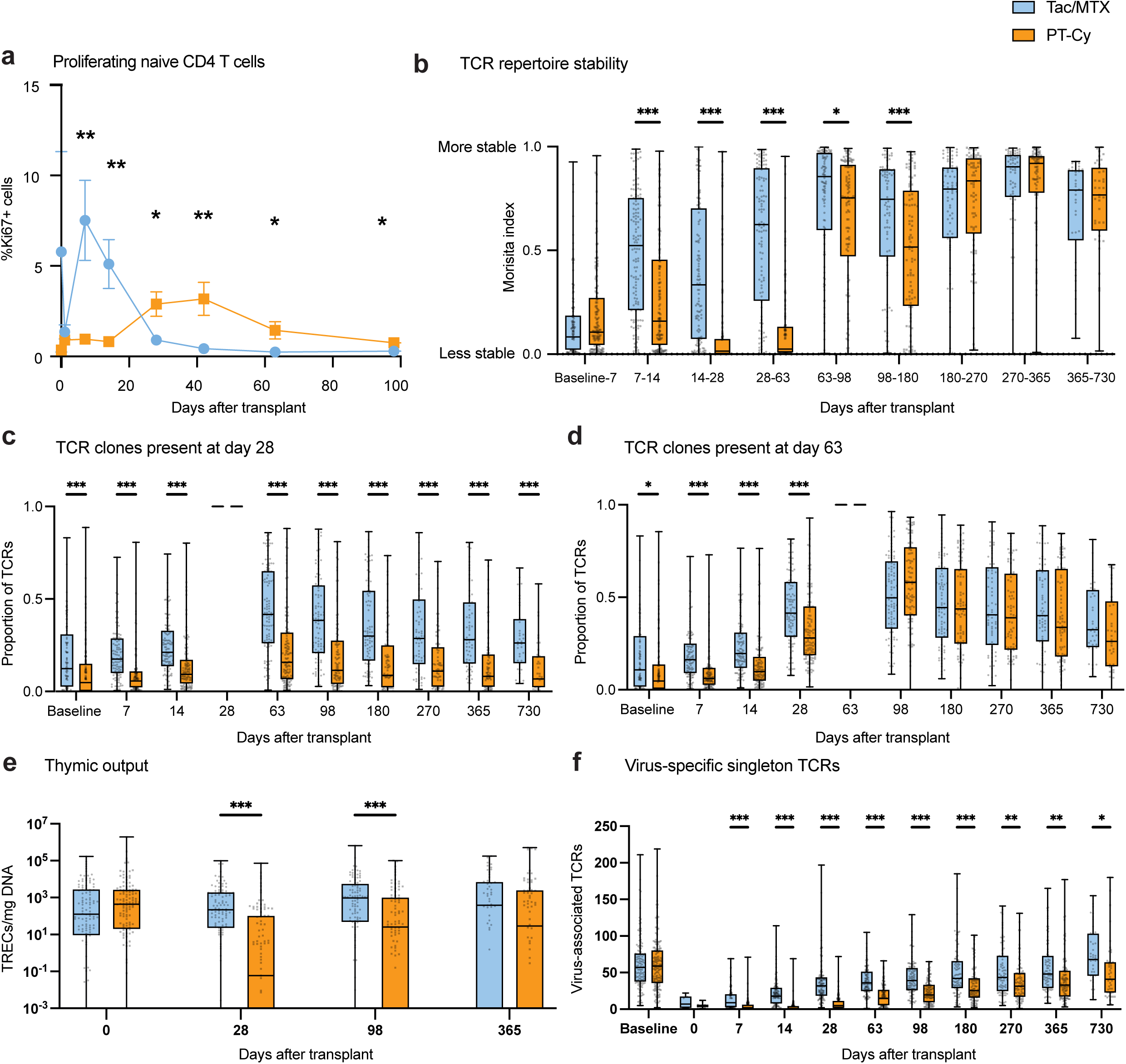
Origin of T cells building the post-transplant immune compartment. a, Intracellular cytokine staining followed by flow cytometry was used to assess Ki67 expression in T cells from each subset (N=40 patients). The frequency of proliferating CD4+ naïve T cells is displayed as mean±SEM at each timepoint. Asterisks indicate P values for the comparison between arms at each timepoint using mixed effects models for repeated measures testing with Fisher’s least significant difference posttests at each timepoint, * P<0.05, ** P<0.01, *** P<0.001. **b,** The Morisita Index, a measure of repertoire stability/turnover, was calculated for each pair of timepoints listed; higher values indicate a more stable TCR repertoire (N=324 patients). TCR clones present at each of Day 28 (Panel **c**) and Day 63 (Panel **d**) were plotted at all timepoints (N=264 and 238 patients, respectively). **e**, Thymic output measured by quantification of T cell receptor excision circles (TREC) from genomic DNA is shown as box and whisker plots stratified by study arm (N=291 patients). Values were compared between arms at each timepoint using multiple Mann-Whitney U tests. **f,** The number of singleton TCRs associated with cytomegalovirus, Epstein-Barr virus, SARS-CoV-2, herpes simplex virus-1, herpes simplex virus-2 and parvovirus are displayed (N=324 patients). Data are represented as mean ± SEM and comparison between groups made by Mann-Whitney U test. Asterisks indicate P values for these comparisons, * P<0.05, ** P<0.01, *** P<0.001.

The timing of the bottleneck release after PT-Cy was measured through a sum frequency analysis of Day 28 (**Figure 5c)** and Day 63 (**Figure 5d**) clones. While few Day 28 TCR clones were present long-term after PT-Cy, clones that emerge by Day 63 persist, consistent with a bottleneck release between these two timepoints. Together these data demonstrate that, while the total number of T cells after Day 63 is similar between Tac/MTX and PT-Cy, the T cells in PT-Cy originate from a smaller founder population, and expand later, leading to a long-term deficit in T-cell diversity versus Tac/MTX.

Recent thymic emigrants (RTEs) are critical for the recovery of post-transplant T-cell immunity. We measured the number of T-cell receptor excision circles (TRECs)^24^ in PT-Cy versus Tac/MTX patients at 4 timepoints (pre-transplant, Days +28, +98 and +365) to compare the contributions of RTEs to post-transplant TCR diversity. As expected, pre-transplant TREC numbers were similar between arms, but PT-Cy patients demonstrated significantly fewer TRECs on Days +28 and +98, with recovery by 1-year post-transplant. This is consistent with cyclophosphamide causing early post-HCT thymic injury, thus further contributing to reduced post-transplant TCR diversity (**Figure 5e)**.

### Functional effect of reduced T cell diversity on antiviral immunity

We next asked whether the constrained TCR repertoire after PT-Cy limited the extent of virus-specific T cells. TCR classifiers are increasingly available for viral infections, and were used in this study to identify clones associated with cytomegalovirus (CMV), Epstein-Barr Virus (EBV), SARS-CoV-2, herpes simplex virus (HSV)-1, HSV-2 and parvovirus. The total number of virus-specific singleton TCRs was significantly lower in PT-Cy-treated patients, starting early post-transplant and persisting at least 2 years (**Figure 5f**). Similar trends were observed for each virus individually (**Extended Data Figure 8**).

## Discussion

We present the findings of a multicenter longitudinal analysis of T-cell composition in HCT patients allocated to one of two prevailing GVHD prophylaxis strategies. This analysis included evaluation of the T-cell repertoire at a depth and scale not previously achieved.^25–30^ We discovered major differences in T-cell dynamics depending on the type of GVHD prophylaxis regimen, including a significant decrease in TCR diversity with PT-Cy, which was present as early as 7 days post-HCT and persisted for >2 years. TCR diversity was associated with long-term rates of both cGVHD and infection, independent of treatment arm, with PT-Cy more likely to associate with low TCR diversity than Tac/MTX, likely explaining its more effective control of cGVHD and improved GRFS, and its association with increased Grade 2 (moderate) infections. PT-Cy patients demonstrated early depletion of all T-cell populations, as well as deficits in thymic output, with subsequent T-cell expansion from a more limited pool of T-cell clones. This smaller number of T-cell clones led to long-term constraints on TCR repertoire diversity. This is consistent with a T-cell bottleneck effect^20–22^ with PT-Cy.

Prior studies of post-HCT immune reconstitution have relied on mouse models or single-regimen, uncontrolled human observational cohorts.^31–38^ In particular, the biology of PT-Cy has not previously been studied in humans in a randomized, comparative fashion. This mechanistic study linked to a large clinical trial enabled the analysis of over 2,000 samples, providing a sample set of sufficient magnitude to overcome the variability inherent in human data,^39^ particularly during immune reconstitution, when multiple events occur simultaneously.

Furthermore, while retrospective work has compared the composition of T cell populations across transplant approaches,^38^ this study analyzes the actual diversity of those T cell subsets, linked to cell phenotype, substantially deepening our understanding of the impact of key GVHD prophylactic regimens on post-transplant T cell immunity.

The results of this study elucidate the immunologic trade-offs associated with different GVHD prophylaxis approaches. PT-Cy led to pan-T cell depletion and a long-lasting decrease in overall T-cell diversity compared to transplant patients treated with Tac/MTX, and to transplant donors. This functionally smaller T-cell pool likely mitigates GVHD by limiting the number of T cells that mediate tissue damage.^40–43^ At the same time, however, this relative T-cell diversity deficit also leads to fewer T cells that could be available to respond to pathogen-associated antigens. It is notable that other than an increase in grade 2 (moderate) infections, the decreased TCR diversity with PT-Cy did not produce major clinical costs, though also did not result in significantly improved overall survival. The opposing effects of decreased TCR diversity may contribute to the lack of significant improvement in overall survival despite the substantial reduction in GVHD with PT-Cy.

The increase in grade 2 (moderate) infections with PT-Cy compared to Tac/MTX was primarily due to bacterial infections early post-transplant.^15^ This observation highlights the often-overlooked role that T cells play in protective immunity against bacteria,^44,45^ especially for tissue-based protection against GI tract bacterial translocation. With respect to viral infections, PT-Cy has previously been associated with cytomegalovirus reactivation and disease,^46^ but the risk of this viral infection (and other viral infections) during this trial was not different between arms, despite decreases in viral-associated T cells with PT-Cy. CMV may have been lessened by the wide use of the anti-CMV drug letermovir.^47^ As such, PT-Cy may result in different infectious risks in places where letermovir use is less common. This analysis was not powered to detect differences in TCR diversity stratified by type of infection, and this represents an important area of future investigation.

The observation that the decreased TCR diversity with PT-Cy did not lead to increased relapses is notable, and suggests that for the reduced-intensity unrelated-donor HCT studied in the BMT CTN 1703/1801 trials, differences in TCR diversity alone do not drive recurrent malignancy.

One hypothesis for a positive impact of PT-Cy on relapse prevention after reduced-intensity HCT relates to the increased exposure to cyclophosphamide in these patients, which could have anti-leukemic effects. Reduced exposure to adjunct immunosuppression in PT-Cy patients by controlling GVHD could also better support graft-versus-leukemia effects. Expanding the analysis of the impact of PT-Cy on transplant outcomes for all conditioning intensities and all leukemia molecular subtypes is merited.

This analysis revealed that TCR diversity early (within 14 days) after HCT predicts key transplant outcomes, raising the potential for early identification of at-risk patients, whether treated with Tac/MTX or PT-Cy. Modification of treatment through, for example, reduction in immunosuppression or augmentation in infection prophylaxis, could be considered. Importantly, our observation of decreased TREC output in the first 100 days post-HCT in patients receiving PT-Cy is in agreement with studies in model organisms documenting direct cyclophosphamide-mediated thymic damage^48^, and suggests that approaches to support thymic recovery may warrant evaluation in PT-Cy-treated patients.

Together, these data provide novel insights into the biologic underpinnings of GVHD and immune reconstitution after HCT and begin to unravel the essential paradox of allogenic HCT: how to balance the hazard of robust donor T-cell number, diversity and function (GVHD) against their potential beneficial effects (engraftment, reductions in infections, and prevention of leukemia relapse). Deconstructing these biologic phenomena will help to guide future innovations in HCT.

## Methods

### Trial design

This study was a companion to the randomized, Phase 3 multicenter BMT CTN 1703 trial.^15^ Key objectives included comparing the diversity of the T-cell receptor (TCR) repertoire after transplant with Tac/MTX versus PT-Cy, and identifying the effects of T-cell diversity on clinical outcomes. Inclusion and exclusion criteria were the same as the paired BMT CTN 1703 trial. Co-enrollment on BMT CTN 1801 was planned for at least 300 patients (70% of the 428-patient enrollment goal for BMT CTN 1703). Informed consent was obtained from all participants. The study protocol was approved by the Institutional Review Board of the National Marrow Donor Program. Submitted on behalf of the Blood and Marrow Transplant Clinical Trials Network (BMT CTN).

### Biologic Samples

Samples were obtained before preparative conditioning (between 7-14 days before transplant), prior to graft infusion (the day prior to or the day of infusion), weekly through the first 84 days after transplant and then on days 98, 180, 270, 365 and 730 after transplant. After transplant infusion, empty stem cell graft bags or syringes were also obtained and shipped to a central reference laboratory for graft cell isolation. Peripheral blood samples were shipped by each site to the central BMT CTN Repository operated by the NMDP where peripheral blood mononuclear cells, whole blood and plasma were isolated and frozen in separate aliquots from each timepoint.

### T-cell receptor variable beta chain sequencing

Immunosequencing of the CDR3 regions of human TCRβ chains was performed using Adaptive Immunosequencing (Adaptive Biotechnologies, Seattle, WA). Extracted genomic DNA was amplified in a bias-controlled multiplex PCR, followed by high-throughput sequencing. Sequences were collapsed and filtered to identify and quantitate the absolute abundance of each unique TCRβ CDR3 region for further analysis as previously described.^17,18,49^

### Multiparameter flow cytometry

Samples for flow cytometry were selected from patients without chronic GVHD or relapse, to help ensure their reflection of the immunosuppressive strategies used, rather than immune-mediated transplant outcomes. The overall similarity of immune reconstitution in the flow cytometry cohort compared to all BMT CTN 1801 patients was assessed through longitudinal analysis of total lymphocyte counts, which were similar between groups (**Extended Data Figure 5a-b**). PBMCs cryopreserved at-150C were thawed slowly into prewarmed (37C) media (RPMI + 10% FBS): 1mL of frozen cells was added dropwise to 1mL of media and serially diluted 1:2 with media until a final volume of 32mL. Cells were strained through a 100um cell strainer to remove clumps, centrifuged at 350g for 10min at 23C, and resuspended in 1mL of media for counting. Viable cells were counted with a Cellometer Auto 2000 Cell Viability Counter (Nexcelom). All cells were stained with LIVE/DEAD Fixable Aqua Dead Cell Stain Kit (ThermoFisher), after which cells were divided in half between two antibody panels: 16-color panel for characterizing T cells and a 14-color panel for characterizing B and NK cells. Flow cytometry data were acquired within 48 hours of staining using a LSRFortessa flow cytometer (BD Biosciences) and analyzed using FlowJo (v10.0.0). T cell subsets were defined as follows: T_CM_ (CCR7+ CD45RA-), T_EM_ (CCR7-CD45RA-), T_EMRA_ (CCR7-CD45RA+), T_N_ (CCR7+ CD45RA+ CD95-), T_SCM_ (CCR7+ CD45RA+ CD95+), T_reg_ (CD4+ CD25+ CD127lo), γδ-T cells (CD3+ γδTCR+). NK cells were defined as CD3-NKp46+. B cells were defined as CD3-CD19+. Antibodies used were the following: CD3-BUV395 (BD Biosciences), CD4-BV786 (BD Biosciences), CD5-BV786 (BD Biosciences), CD8-APC-Cy7 (BD Biosciences), CD14-BUV661 (Invitrogen), CD16-APC (BD Biosciences), CD19-PE (Abcam), CD20-BUV661 (BD Biosciences), CD21-BUV737 (BD Biosciences), CD24-APC-Cy7 (BioLegend), CD25-BB515 (BD Biosciences), CD27-BV421 (BD Biosciences), CD28-BUV737 (BD Biosciences), CD38-BV605 (BD Biosciences), CD45RA-PE-Cy7 (BioLegend), CD56-BB515 (BD Biosciences), CD69-BUV496 (BD Biosciences), CD95-AF700 (BioLegend), CD127-PE (BD Biosciences), PD-1-BV711 (BD Biosciences), CD335-PE-Cy7 (BioLegend), CCR7-BV421 (BD Biosciences), gdTCR-BB700 (BD Biosciences), GranzymeB-APC (BioLegend), HLA-DR-BUV496 (BD Biosciences), IgD-BV711 (BD Biosciences), Ki67-PerCP-Cy5.5 (BD Biosciences).

Representative gating is shown in **Supplementary Figure 1**.

### Single-cell RNA sequencing

A subset of the PBMCs from patients analyzed by flow cytometry who had PBMCs frozen from the leftover graft bag/syringe also underwent scRNA-and TCR-Seq. These PBMCs were enriched for CD3+ T cells through MACS using the Miltenyi human Pan T cell Isolation Kit (Miltenyi Biotec). Approximately 16,500 cells per sample were loaded onto a 10X Chromium Instrument (10x Genomics) according to manufacturer’s instructions. GEX libraries were generated using Chromium Next GEM Chip K Single Cell Kit v2 (10X Genomics) and normalized to 12nM; VDJ libraries were generated using Chromium Next GEM Single Cell 5’ Kit v2 (10X Genomics) and normalized to 3nM. Pooled GEX and VDJ libraries were sequenced on Illumina NovaSeq S4 at the Broad Institute.

Analysis of the 5’ scRNA + TCR data was performed as follows: FASTQ files were aligned with Cell Ranger v7.1.0 using GRCh38 as the reference genome, then imported into Python v3.10.12. Droplets were filtered for protein-coding, TR_C_, and IG_C_ genes per annotations from Ensembl v112. We then performed QC to analyze only cells with a mitochondrial gene expression of less than 10%, between 700 and 6,000 unique genes, and between 1,000 and 25,000 transcripts. We further filtered the genes to those expressed in at least 10 cells and then selected the 7,500 most variable genes (running Scanpy v1.10.2) plus *PTPRC* and *CD3E*. To filter for T cells, we performed initial clustering with Scanpy and removed clusters with low *CD3E* expression. We then performed clustering using an encoding generated with scvi-tools v1.1.5 and removed another 2 clusters low in *CD3E* expression. For all downstream analyses, we focused on D28 and D365 samples. We manually annotated separate D28 and D365 CD4 and CD8 clusters using a list of T cell marker genes and studying the list of differentially expressed genes within each cluster after batch correcting by patient. For TCR analysis, we extracted clonotype groupings generated by Cell Ranger, which were subsequently aligned with Adaptive TRB CDR3 sequences when appropriate.

### T-cell Receptor Excision Circle (TREC) analysis

Genomic DNA extracted for TCR sequencing was subjected to TREC analysis. TREC concentrations were analyzed by real-time quantitative PCR (Bio-Rad) using TaqMan Gene Expression Master Mix (Applied Biosystems). The RT-qPCR reactions were performed in a final volume of 25 μL following the protocol described by Sempowski et al.^50^. Where possible, 10 μL of sample were loaded into each well. In each reaction, the DNA sample was tested in duplicate.

A standard curve was included in each qPCR plate. The number of TRECs/well was normalized to the amount of DNA/well.

### Statistical analysis

TCR diversity was expressed as the inverse Simpson diversity index, as specified in the BMT CTN 1801 Statistical Analysis Plan (SAP). Additional diversity metrics were also explored, as described in Supplemental Methods. Log-normalized diversity values were compared between treatment arms using a mixed-effects model to account for repeated measures with missing values, followed by Fisher’s least significant difference tests at each timepoint. The following additional variables were also tested as candidate covariates for stepwise inclusion in a multivariate model, as specified in the SAP, but none reached the inclusion threshold of P <0.05 (**Supplementary Table 7**): gender, race, age, primary disease, conditioning regimen, disease risk index (DRI), and degree of HLA match. Associations between TCR diversity and clinical outcomes were assessed as described below. Overall survival, relapse and non-relapse mortality were specified in the SAP; cGVHD and grade 2-3 infection were examined due to their identification in BMT CTN 1703 as key discriminators between the Tac/MTX and PT-Cy arms, results that became available after the SAP was developed. Landmark analyses were performed using day 98 and day 180 diversity values, as specified in the SAP. Exploratory analyses stratifying patients by Day 14 diversity were also performed as this timepoint demonstrated the largest differences in TCR diversity between the Tac/MTX and PT-Cy arms (**Supplementary Table 3**).

### Diversity Metrics

The number of unique TCR clonotypes identified in a sample is defined as the richness of that sample. To incorporate information about the evenness of the distribution of different clonotypes, additional metrics are calculated. The inverse Simpson diversity index represents the probability of two TCRs chosen at random from the sample being different; larger values reflect a more even distribution of more clonotypes. Shannon Entropy also incorporates both species richness and evenness, but is more sensitive to the number of clonotypes present while being less sensitive to clonotype dominance.^19^ Singletons are TCRs found exactly once in a sample. The ratio of singletons to richness represents the proportion of unique TCR clonotypes that were found exactly once rather than multiple times in a sample.

### Associations between TCR diversity and clinical outcomes

As specified in the Statistical Analysis Plan, landmark analyses were undertaken at day 98 and 180. To test associations between TCR diversity and clinical outcomes, patients were stratified into high and low diversity groups using the median inverse Simpson diversity value at each specified timepoint, including only patients who had a sample available at those timepoints. The cumulative incidence of each outcome developing subsequent to that timepoint was assessed. Tested outcomes were overall survival, relapse/progression, non-relapse mortality, cGVHD requiring systemic immunosuppression and grade 2-3 (moderate-to-severe) infection. The cGVHD and infection outcomes were not specified in the SAP, but added based on results from the BMT CTN 1703 trial. A day 14 analysis was also added based on results demonstrating this was the most discriminative timepoint for diversity between PT-Cy and Tac/MTX arms. This analysis was not performed using a day 14 landmark, but rather by stratifying all patients based on median day 14 inverse Simpson diversity value, then plotting the cumulative incidence from day 0 of each outcome.

## Data Availability Statement

TCR sequencing data is available through immuneACCESS (DOI: 10.21417/SS2024NM). Single-cell RNA sequencing is available through the National Center for Biotechnology Information Gene Expression Omnibus database (accession GSE285821). Deidentified participant data and the study protocol will be available in the National Heart, Lung, and Blood Institute Biologic Specimen and Data Repository Information Coordinating Center (BioLINCC) (https://biolincc.nhlbi.nih.gov/home/), subject to their standard policies and procedures.

## Extended Data Legends

**Extended Data Figure 1:** A heatmap was constructed for each GVHD prophylaxis arm, with each row indicating a separate patient and each column a different timepoint. A filled box indicates TCR sequencing was performed at that timepoint for that patient.

**Extended Data Figure 2:** Additional TCR diversity metrics are displayed for Tac/MTX versus PT-Cy treated patients, including richness (Panel **a**), singletons (Panel **b**) and the proportion of richness that is singletons (Panel **c**), after computationally down-sampling each sample to 5,000 TCRs (N=324 patients). Asterisks indicate P values for these comparisons, * P<0.05, ** P<0.01, *** P<0.001.

**Extended Data Figure 3:** Singletons were measured at different down-sampling targets ranging from 1,000 to 50,000 TCRs (N=324 patients). Asterisks indicate P values for these comparisons, * P<0.05, ** P<0.01, *** P<0.001.

**Extended Data Figure 4:** Patients with Day 14 samples available (N=281 patients) were dichotomized into high and low diversity groups using the median inverse Simpson diversity index value. The cumulative incidence of chronic GVHD requiring immunosuppression (Panel **a**) and moderate-to-severe infections (Panel **b**) through 1 year after transplant were plotted, with patients in each GVHD prophylaxis arm plotted separately. The cumulative incidence of non-relapse mortality (Panel **c**) and relapse (Panel **d**) were plotted through 1 year after transplant. The 1-year estimates of the cumulative incidence are provided by group along with 95% confidence intervals. Gray’s test was used for comparisons between high and low diversity groups.

Univariate Fine-Gray regression models were also fit for each outcome and set of diversity groups and used to present a subdistribution hazard ratio representing the multiplicative impact on the cumulative incidence of relapse for the lower diversity group relative to the higher diversity group. The competing risk for non-relapse mortality was relapse. All endpoints were censored at 1-year post-transplant or last follow-up, whichever occurred first.

**Extended Data Figure 5: a,** The absolute lymphocyte count was measured clinically for each enrolled patient at each timepoint (N=324 patients). Data are presented as mean±SEM, and values were compared between arms at each timepoint using mixed effects models for repeated measures testing with Fisher’s least significant difference posttests at each timepoint. Asterisks indicate P values for these comparisons, * P<0.05, ** P<0.01, *** P<0.001. **b,** The absolute lymphocyte counts for only those patients (N=40) selected for flow cytometry on frozen peripheral blood mononuclear cells are displayed. **c to g,** Multiparameter flow cytometry was used to identify the absolute number of lymphocytes in each indicated cell subset (N=40 patients).

**Extended Data Figure 6: a to h**, Multiparameter flow cytometry was used to identify the absolute number of lymphocytes in each indicated cell subset (N=40 patients). Data are presented as mean±SEM, and values were compared between arms at each timepoint using mixed effects models for repeated measures testing with Fisher’s least significant difference posttests at each timepoint. Asterisks indicate P values for these comparisons, * P<0.05, ** P<0.01, *** P<0.001.

**Extended Data Figure 7: a**, Gene marker plot for main T cell phenotypes defined for the single cell RNA sequencing data (for day 28 and day 365 T cells, N=13 patients). **b,** UMAP highlighting all T cells with a detectable TCR in single cell RNA sequencing data (gray) and those TCRs detected in both single cell and bulk sequencing platforms (blue dots).

**Extended Data Figure 8:** The number of singleton TCRs associated with cytomegalovirus (**a**), Epstein-Barr virus (**b**), SARS-CoV-2 (**c**), herpes simplex virus-1 (**d**), herpes simplex virus-2 (**e**) and parvovirus (**f**) are displayed (N=324 patients). Data are represented as mean ± SEM and comparison between groups made by Mann-Whitney U test. Asterisks indicate P values for these comparisons, * P<0.05, ** P<0.01, *** P<0.001.

**Extended Data Table 1:** Baseline and transplant characteristics of patients enrolled in this study (BMT CTN 1801) were compared using Kruskal-Wallis tests to those enrolled in the parent trial (BMT CTN 1703) who did not co-enroll in this study.

**Extended Data Table 2:** The inverse Simpson diversity index at day 14 post-transplant was used to stratify patients into high (above the median) and low (below the median) diversity groups. The 1-year estimates of cumulative incidence for each outcome is provided along with 95% confidence intervals. Gray’s test was used for comparisons between high and low diversity groups. Univariate Fine-Gray regression models were also fit for each outcome and set of diversity groups and used to present a subdistribution hazard ratio representing the multiplicative impact on the cumulative incidence of each outcome for the lower diversity group relative to the higher diversity group.

## Acknowledgments

Support for this study was provided by grants #U10HL069294 and #U24HL138660 to the Blood and Marrow Transplant Clinical Trials Network from the National Heart, Lung, and Blood Institute and the National Cancer Institute. The content is solely the responsibility of the authors and does not necessarily represent the official views of the NIH. Support was also provided by the Biostatistics Shared Resource at the Medical College of Wisconsin Cancer Center. This manuscript was prepared using BMT CTN 1703/1801 Research Materials obtained from the BMT CTN Repository operated by the NMDP and does not necessarily reflect the opinions or views of the BMT CTN 1703/1801 protocol team, the BMT CTN, the NHLBI, or NCI.

