## Extended Data Figures for "Graft-versus-host disease prophylaxis shapes T cell biology and immune reconstitution after hematopoietic cell transplant"

**Extended Data Figure 1: Heatmap of TCR-sequencing samples by patient and GVHD prophylaxis arm**

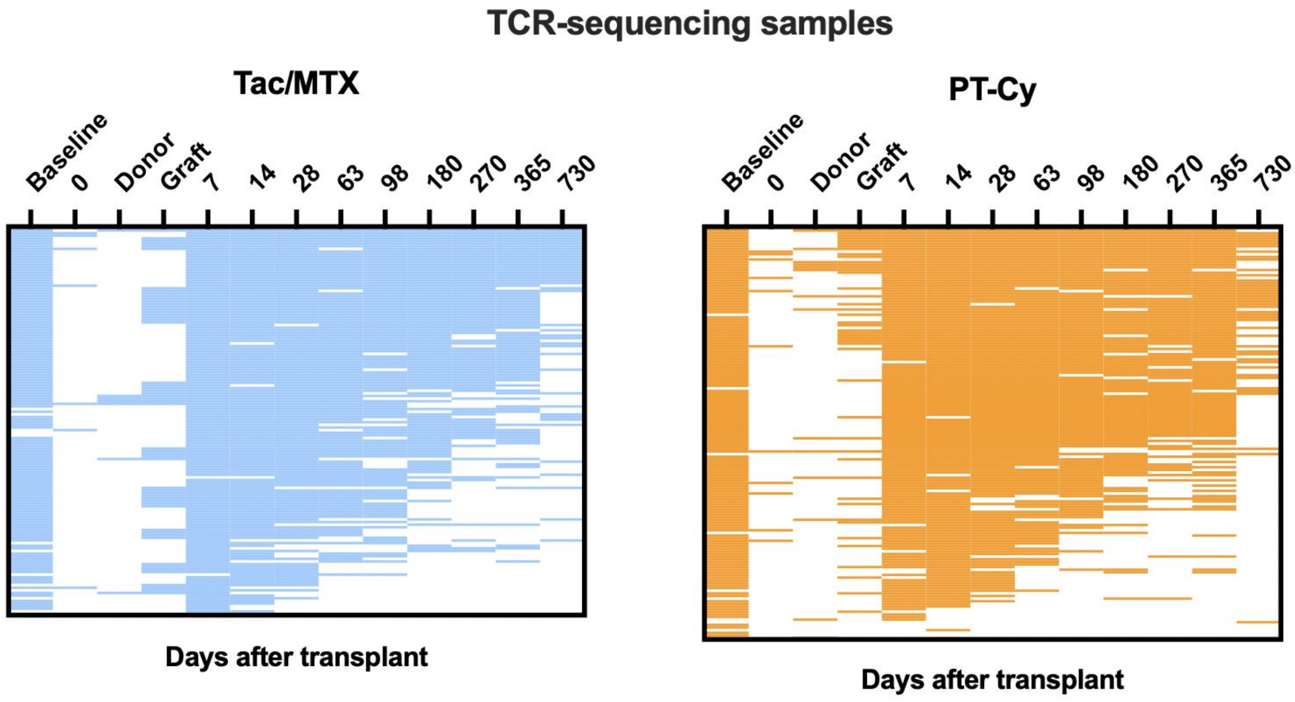

**Extended Data Figure 2: Additional TCR diversity metrics**

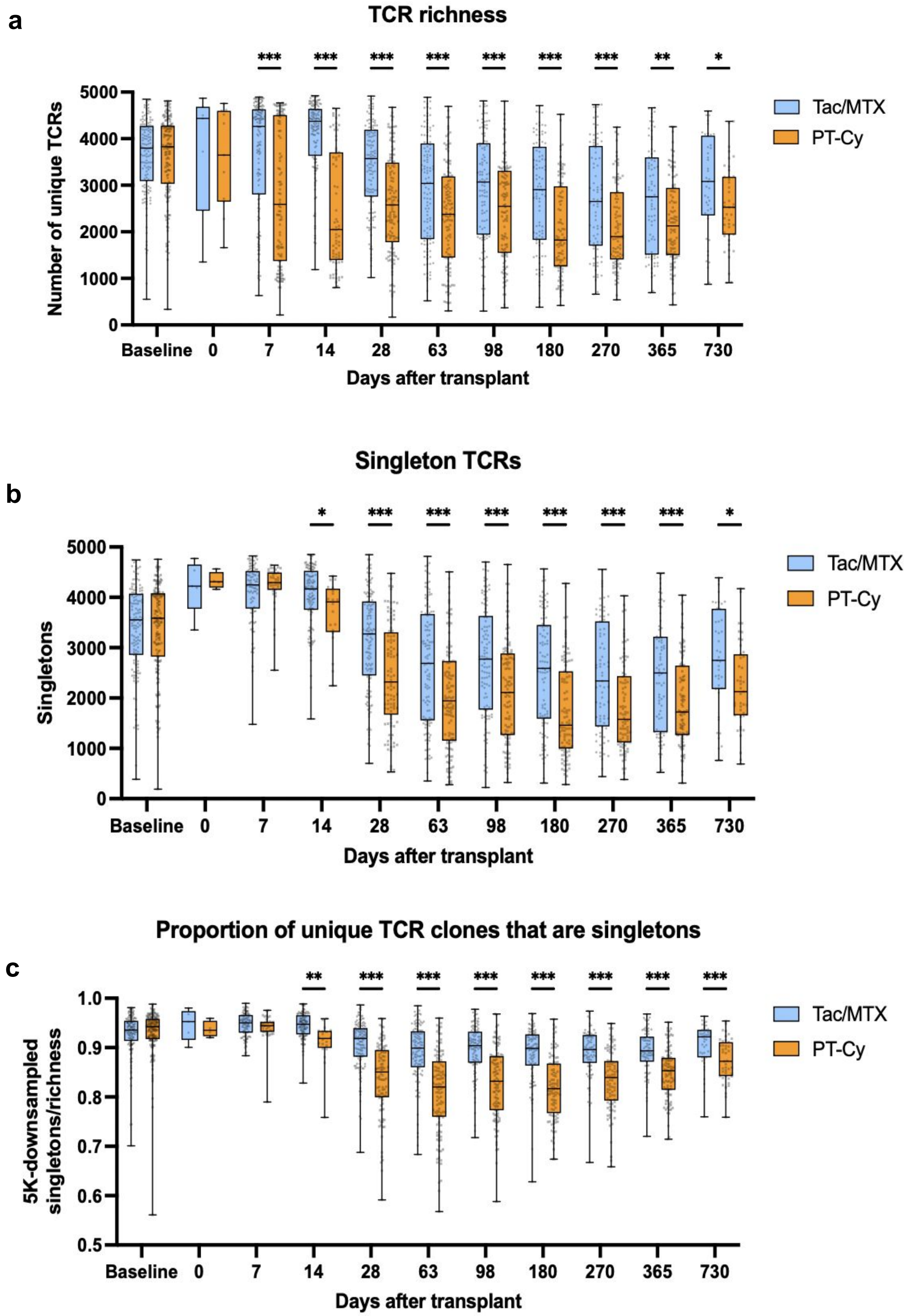

**Extended Data Figure 3: Singleton TCR measurements are robust to different down-sampling targets**

**a**

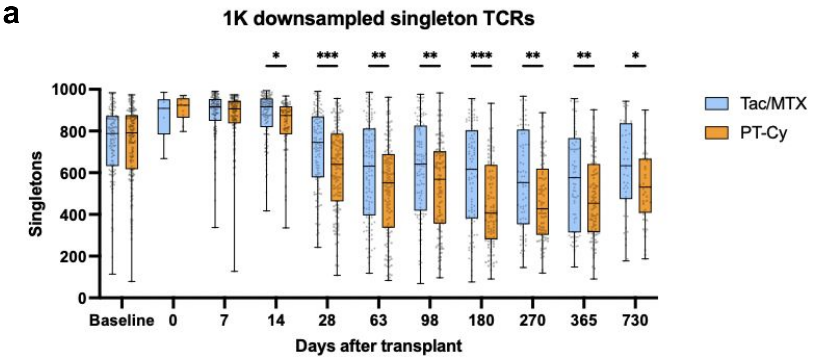


**b**

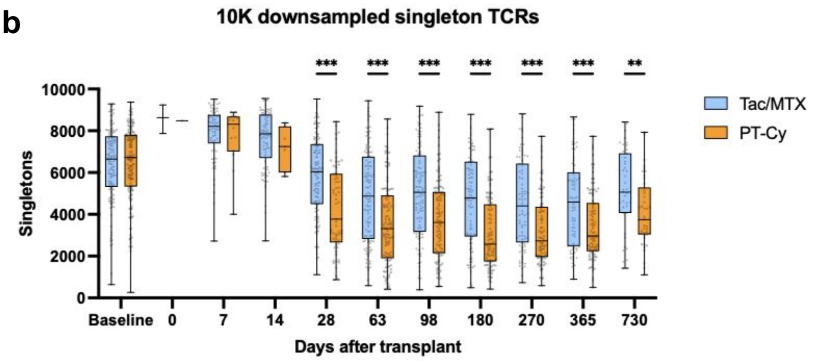


**c**

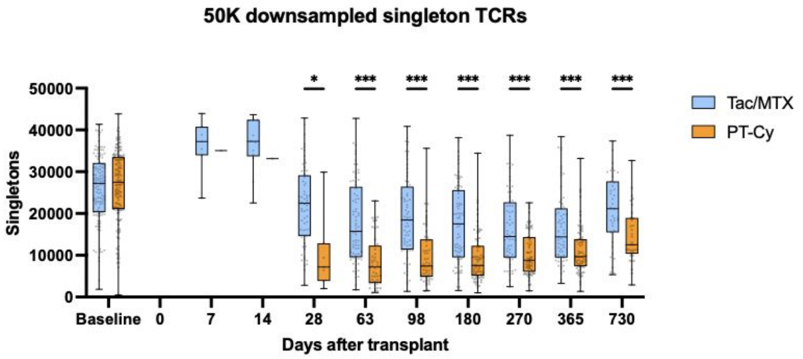

### Extended Data Figure 4: Association of Day 14 inverse Simpson diversity with additional outcomes

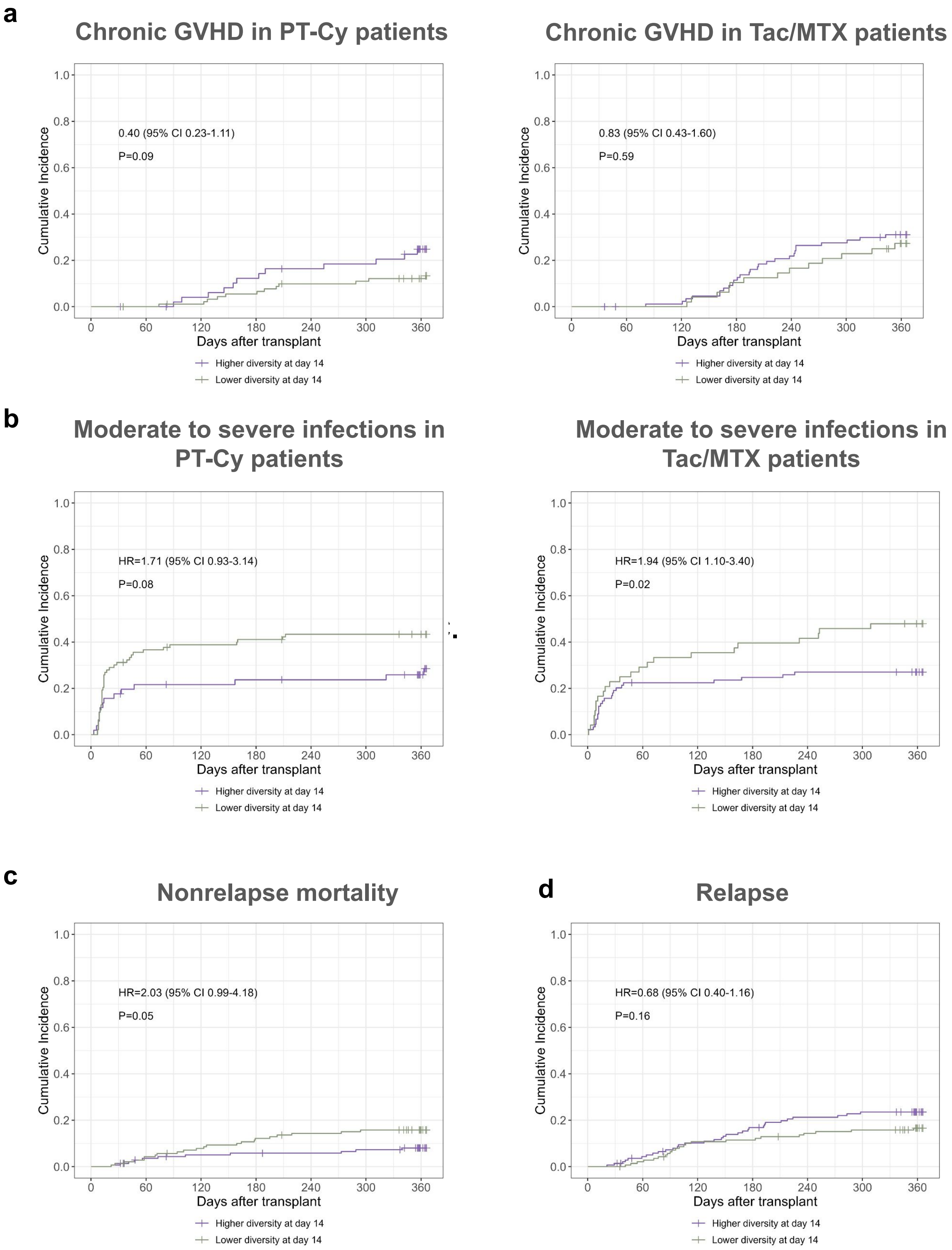

**Extended Data Figure 5: Flow cytometry analysis of immune reconstitution according to GVHD prophylaxis**

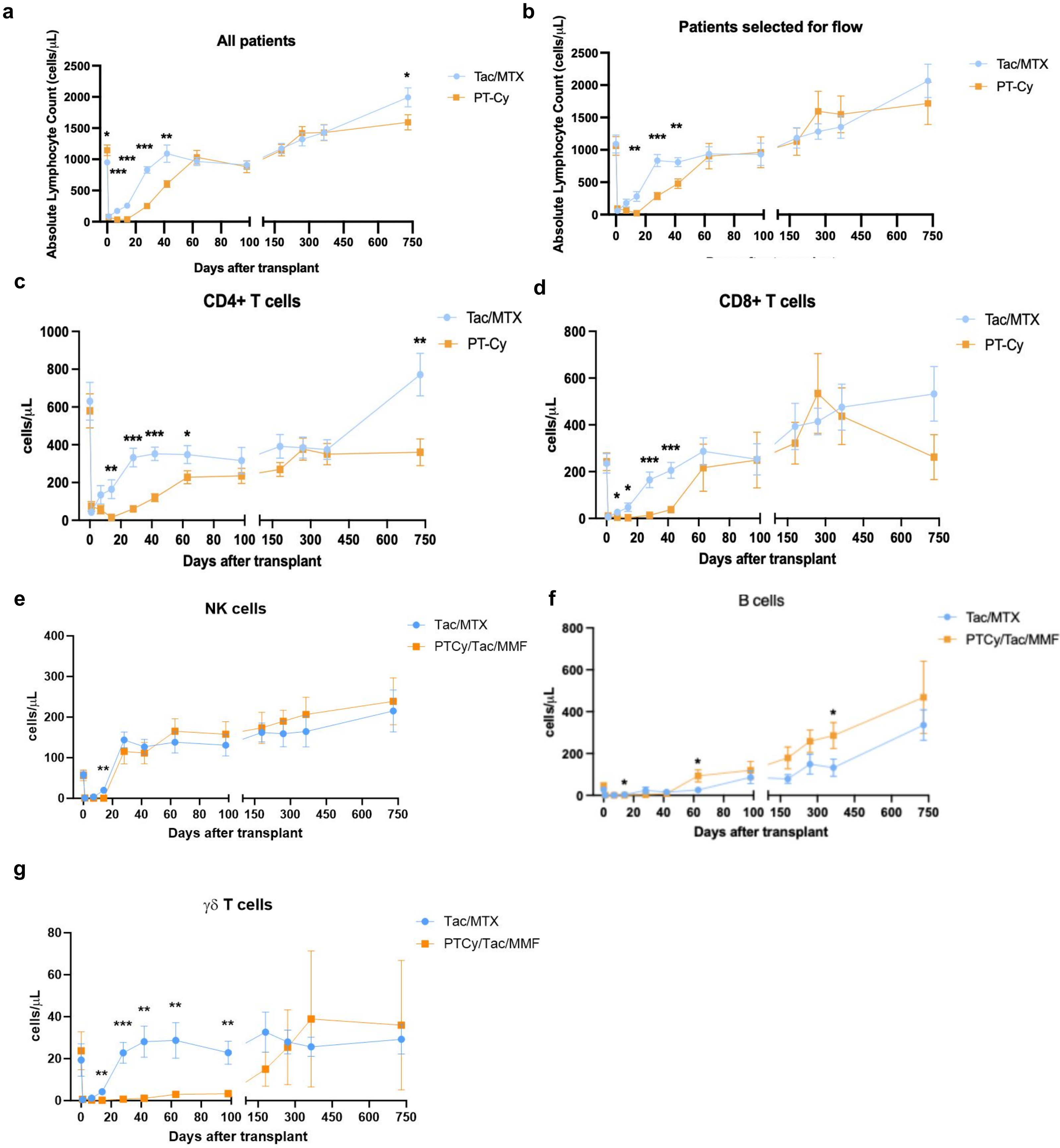

#### Extended Data Figure 6: Flow cytometry analysis of CD4+ and CD8+ subsets

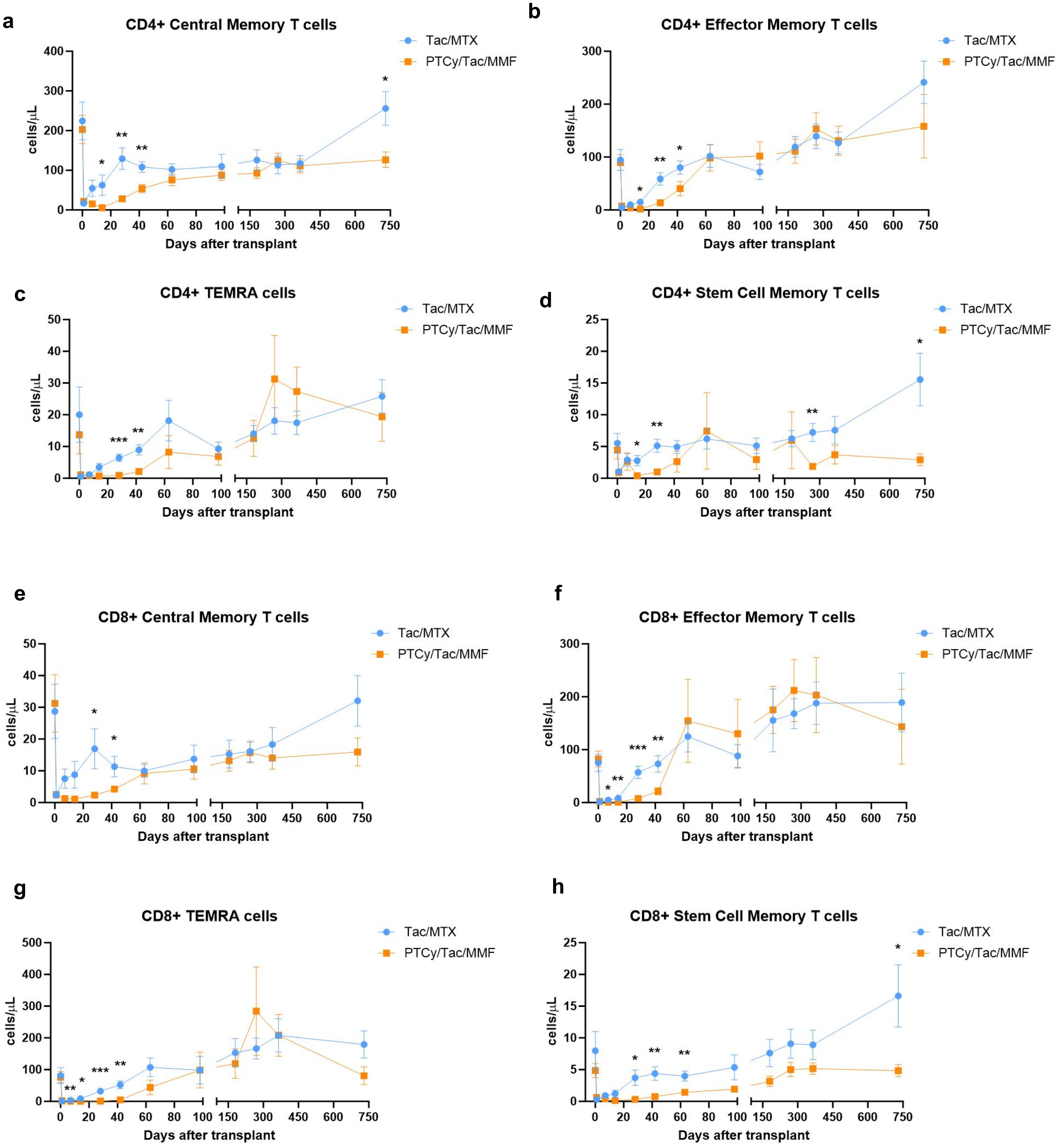

**Extended Data Figure 7: Single-cell RNA-sequencing analysis of T cell phenotype**

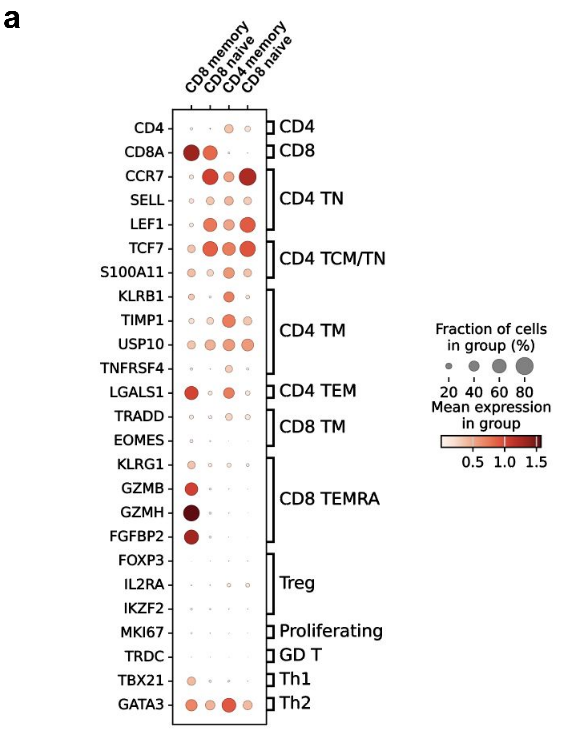


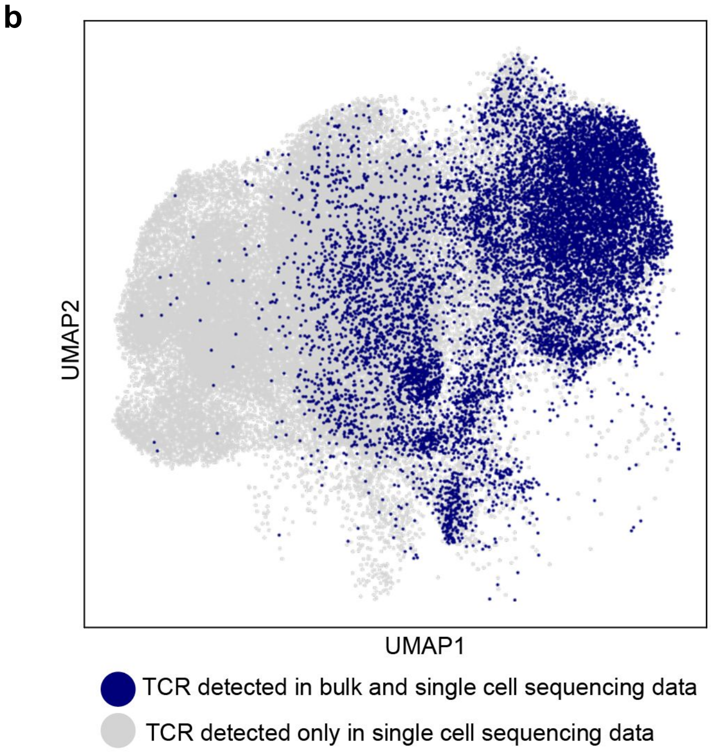

### Extended Data Figure 8: TCR singletons associated with individual viruses

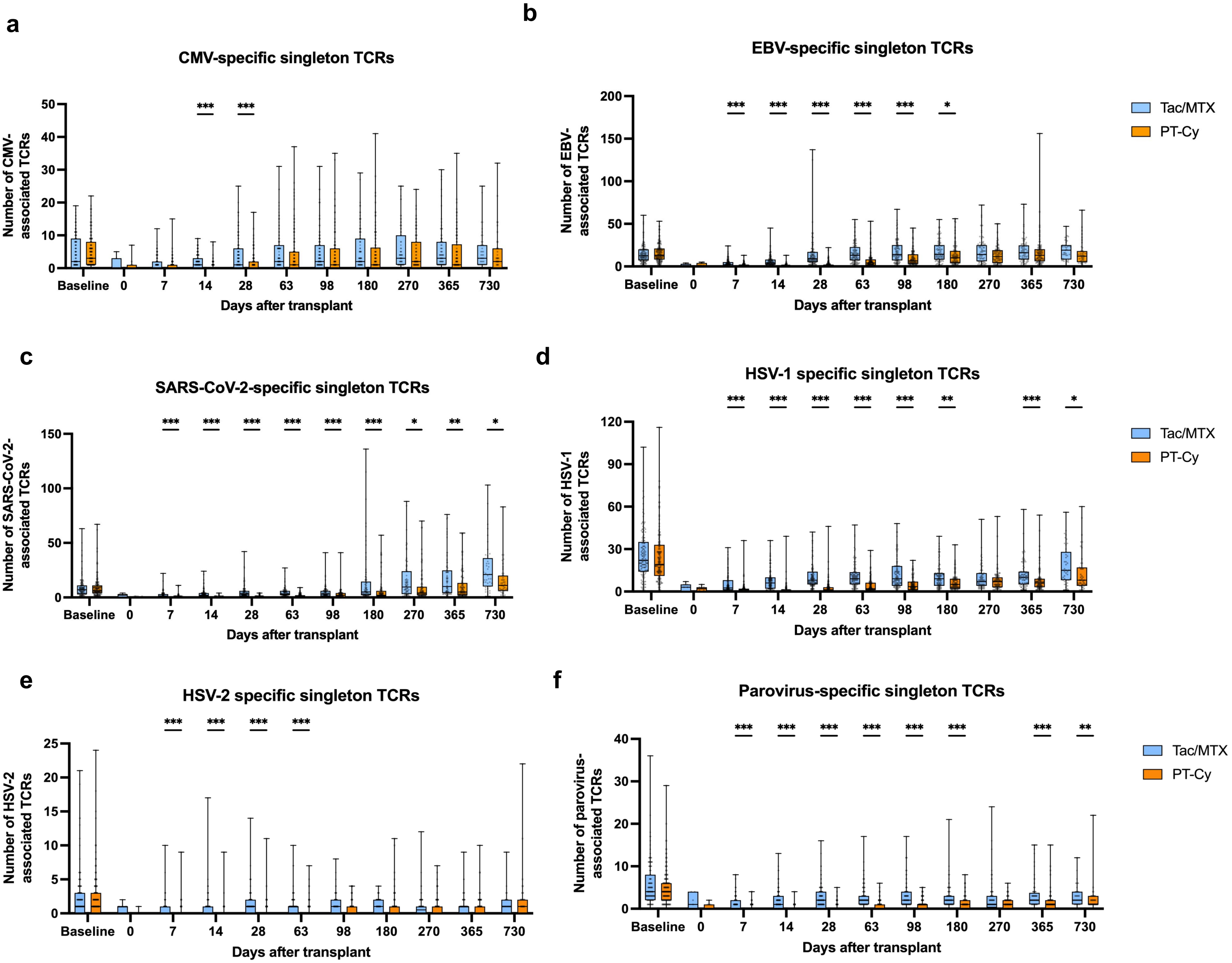

**Extended Data Table 1**

| Characteristics of parent study patients co-enrolled on this study | | | | | |
|---|---|---|---|---|---|
| | Co-enrolled in BMT CTN 1801 trial (this study) | | Not co-enrolled in BMT CTN 1801 trial (only enrolled on parent study) | | |
| **Characteristic** | **Tac/MTX group (N=159)** | **PT-Cy group (N=165)** | **Tac/MTX group (N=58)** | **PT-Cy group (N=49)** | **P value for comparison among groups** |
| Age – mean in yrs ± SD | 64.6 ± 8.2 | 64.4 ± 8.2 | 64.2 ± 10.5 | 63.4 ± 9.4 | 0.85 |
| Male sex – no. (%) | 91 (57.2) | 99 (60.0) | 35 (60.3) | 35 (71.4) | 0.10 |
| Race or ethnic group – no. (%) | | | | | |
| Hispanic or Latinx ethnic group | | | | | 0.78 |
| Hispanic or Latinx | 15 (9.4) | 7 (4.2) | 7 (12.1) | 2 (4.1) | |
| Not Hispanic or Latinx | 143 (89.9) | 156 (94.5) | 48 (82.8) | 47 (95.9) | |
| Not reported or unknown | 1 (0.6) | 2 (1.2) | 3 (5.2) | 0 | |
| American Indian or Alaska Native | 1 (0.6) | 0 | 0 | 0 | 0.48 |
| Asian | 3 (1.9) | 8 (4.8) | 1 (1.7) | 2 (4.1) | |
| Black | 4 (2.5) | 8 (4.8) | 1 (1.7) | 0 | |
| Native Hawaiian or Pacific Islander | 0 | 0 | 0 | 0 | |
| White | 142 (89.3) | 139 (84.2) | 51 (87.9) | 47 (95.9) | |
| Multiple | 0 | 0 | 1 (1.7) | 0 | |
| Unknown | 9 (5.7) | 10 (6.1) | 4 (6.9) | 0 | |
| Primary disease – no. (%) | | | | | 0.22 |
| Acute lymphoblastic leukemia | 18 (11.3) | 9 (5.5) | 9 (15.5) | 3 (6.1) | |
| Acute myeloid leukemia | 74 (46.5) | 83 (50.3) | 26 (44.8) | 24 (49.0) | |
| Myelodysplastic syndrome | 47 (29.6) | 53 (32.1) | 18 (31.0) | 10 (20.4) | |
| Other | 20 (12.6) | 20 (12.1) | 3 (5.2) | 10 (20.4) | |
| Donor type and HLA matching – no. (%) | | | | | 0.49 |
| Related donor 6/6 | 49 (30.8) | 46 (27.9) | 19 (32.8) | 14 (28.6) | |
| Unrelated donor 8/8 | 104 (65.4) | 114 (69.1) | 37 (63.8) | 33 (67.3) | |
| Unrelated donor 7/8 | 6 (3.8) | 5 (3.0) | 2 (3.4) | 2 (4.1) | |
| CMV status (Donor/recipient) – no. (%) | | | | | 0.15 |
| -/- | 52 (32.7) | 43 (26.1) | 18 (31.0) | 16 (32.7) | |
| +/- | 24 (15.1) | 15 (9.1) | 8 (13.8) | 5 (10.2) | |
| -/+ | 28 (17.6) | 54 (32.7) | 12 (20.7) | 13 (26.5) | |
| +/+ | 52 (32.7) | 49 (29.7) | 19 (32.8) | 12 (24.5) | |
| Missing/unknown | 3 (1.9) | 4 (2.4) | 1 (1.7) | 3 (6.1) | |

**Extended Data Table 2**

| Transplant outcomes stratified by day 14 TCR inverse Simpson diversity index | | | | |
|---|---|---|---|---|
| | Estimate at 1 year, % (95% confidence intervals) | | | |
| **Endpoint** | **Higher diversity at day 14 (N=140)** | **Lower diversity at day 14 (N=141)** | **P value** | **Subdistribution Hazard Ratio (95% confidence intervals)** |
| Cumulative incidence of chronic GVHD requiring immunosuppression | 28.9 (21.5—36.7) | 18.2 (12.2—25.1) | 0.035 | 0.59 (0.36—0.97) |
| Cumulative incidence of Grade 2 or 3 Infection | 27.5 (20.3—35.1) | 45.0 (36.6—53.1) | 0.004 | 1.82 (1.22—2.72) |
| Infection-free survival | 61.5 (53.9—70.2) | 46.9 (39.3—56.0) | 0.01 | 1.56 (1.09—2.22) |
| Overall survival | 79.3 (72.8—86.5) | 74.7 (67.8—82.3) | 0.34 | 1.27 (0.77—2.09) |
| Cumulative incidence of non-relapse mortality | 8.1 (4.3—13.4) | 15.8 (10.3—22.4) | 0.05 | 2.03 (0.99—4.18) |
| Cumulative incidence of Grade III or IV acute GVHD | 11.6 (7.0—17.7) | 13.6 (8.5—19.8) | 0.61 | 1.19 (0.61—2.3) |
| Cumulative incidence of relapse | 23.5 (16.8—31.0) | 16.6 (10.9—23.3) | 0.16 | 0.68 (0.40—1.16) |
