## Supplemental Information for "Graft-versus-host disease prophylaxis shapes T cell biology and immune reconstitution after hematopoietic cell transplant"

### Supplementary Information

#### Table of Contents


Supplementary Table and Data Legends Page 2
Supplementary Table 1 Page 4
Supplementary Table 2 Page 5
Supplementary Table 3 Page 6
Supplementary Table 4 Page 7
Supplementary Table 5 Page 8
Supplementary Table 6 Page 9
Supplementary Table 7 Page 10
Supplementary Figure 1 Page 11

#### Supplementary Table Legends

**Supplementary Table 1:** The number of samples that underwent TCR-seq at each timepoint is displayed, divided by treatment arm.

**Supplementary Table 2:** Baseline and transplant characteristics of patients who were selected for flow cytometry and scRNA-seq analysis

**Supplementary Table 3:** To identify the most discriminative timepoint for inverse Simpson diversity between GVHD prophylaxis arms, the mean diversity index value was calculated for each arm at each timepoint. The ratio of this mean for Tac/MTX to PT-Cy is displayed for each timepoint, with a higher ratio indicating a larger difference between arms.

**Supplementary Table 4:** The inverse Simpson diversity index at day 98 post-transplant was used to stratify patients into high (above the median) and low (below the median) diversity groups. Landmark analyses comparing these high and low diversity groups were performed for the relapse, non-relapse mortality, overall survival, chronic GVHD, and infection endpoints. In these landmark analyses, only patients who were alive and event-free with respect to the outcome in question at the landmark time were included. For the cGVHD endpoint, patients with acute GVHD prior to the landmark time were also excluded, given the use of TCR-modifying adjunct immunosuppressive drugs after an acute GVHD diagnosis. Endpoint intervals start at the landmark time, and span until the event, a competing risk, last follow-up, or 1 year from transplant, whichever occurred first. The results of these landmark analyses are interpretable as a conditional assessment of later events beyond the landmark time for each clinical outcome, among those alive and event-free up through the landmark time. Gray's test was used for comparisons between high and low diversity groups. Univariate Fine-Gray regression models were also fit for each outcome and set of diversity groups and used to present a subdistribution hazard ratio representing the multiplicative impact on the cumulative incidence of each outcome for the lower diversity group relative to the higher diversity group.

**Supplementary Table 5:** The inverse Simpson diversity index at day 180 post-transplant was used to stratify patients into high (above the median) and low (below the median) diversity groups. Landmark analyses comparing these high and low diversity groups were performed for the relapse, non-relapse mortality, overall survival, chronic GVHD, and infection endpoints. In these landmark analyses, only patients who were alive and event-free with respect to the outcome in question at the landmark time were included. For the cGVHD endpoint, patients with acute GVHD prior to the landmark time were also excluded, given the use of TCR-modifying adjunct immunosuppressive drugs after an acute GVHD diagnosis.. Endpoint intervals start at the landmark time, and span until the event, a competing risk, last follow-up, or 1 year from transplant, whichever occurred first. The results of these landmark analyses are interpretable as a conditional assessment of later events beyond the landmark time for each clinical outcome, among those alive and event-free up through the landmark time. Gray's test was used for comparisons between high and low diversity groups. Univariate Fine-Gray regression models were also fit for each outcome and set of diversity groups and used to present a subdistribution hazard ratio representing the multiplicative impact on the cumulative incidence of each outcome for the lower diversity group relative to the higher diversity group.

**Supplementary Table 6:** The inverse Simpson diversity index at day 14 post-transplant was used to stratify patients into high (above the median) and low (below the median) diversity groups. The number of patients from each GVHD prophylaxis regimen who were stratified into each diversity group is displayed.

**Supplementary Table 7:** Potential covariates were tested for inclusion in the mixed effects model comparing inverse Simpson diversity index values for TCR diversity between GVHD prophylaxis arms. Per the trial SAP, covariates were to be included if $P<0.05$.

**Supplementary Figure 1:** Representative gating strategy for each of the two multicolor flow cytometry panels denoting analyzed lymphocyte subsets

**Supplementary Table 1**

| TCR-seq samples sequenced | | |
|---|---|---|
| | Treatment Arm | |
| **Sample source** | **Tac/MTX** | **PT-Cy** |
| **Patient** | | |
| Baseline (pre-conditioning) | 135 | 147 |
| Day 0 (pre-infusion) | 7 | 11 |
| Day 7 | 146 | 145 |
| Day 14 | 137 | 144 |
| Day 28 | 130 | 134 |
| Day 63 | 116 | 122 |
| Day 98 | 107 | 110 |
| Day 180 | 98 | 98 |
| Day 270 | 80 | 88 |
| Day 365 | 80 | 98 |
| Day 730 | 43 | 43 |
| **Donor** | | |
| Graft | 55 | 46 |
| Donor | 8 | 15 |

**Supplementary Table 2**

| Characteristics of patients selected for immunophenotyping | | | |
|---|---|---|---|
| **Characteristic** | **Tac/MTX group (N=20)** | **PT-Cy group (N=20)** | **All patients (N=40)** |
| Age – mean in yrs ± SD | 64.1 ± 10.0 | 62.8 ± 6.0 | 63.4 ± 8.2 |
| Male sex – no. (%) | 12 (60.0) | 10 (50.0) | 22 (55.0) |
| Race or ethnic group – no. (%) | | | |
| Hispanic or Latinx ethnic group | | | |
| Hispanic or Latinx | 1 (5.0) | 1 (5.0) | 2 (95.0) |
| Not Hispanic or Latinx | 19 (95.0) | 19 (95.0) | 38 (95.015) |
| Not reported or unknown | 0 | 0 | 0 |
| American Indian or Alaska Native | 0 | 0 | 0 |
| Asian | 0 | 1 (5.0) | 1 (2.5) |
| Black | 0 | 1 (5.0) | 1 (2.5) |
| Native Hawaiian or Pacific Islander | 0 | 0 | 0 |
| White | 20 (100.0) | 16 (80.0) | 36 (90.0) |
| Multiple | 0 | 0 | 0 |
| Unknown | 0 | 2 (10.0) | 2 (5.0) |
| Primary disease – no. (%) | | | |
| Acute lymphoblastic leukemia | 2 (10.0) | 3 (15.0) | 5 (12.5) |
| Acute myeloid leukemia | 14 (70.0) | 8 (40.0) | 22 (55.0) |
| Myelodysplastic syndrome | 4 (20.0) | 3 (15.0) | 7 (17.5) |
| Other | 0 | 6 (30.0) | 40 (12.3) |
| Donor type and HLA matching – no. (%) | | | |
| Related donor 6/6 | 5 (25.0) | 7 (35.0) | 12 (30.0) |
| Unrelated donor 8/8 | 14 (70.0) | 13 (65.0) | 27 (67.5) |
| Unrelated donor 7/8 | 1 (5.0) | 0 | 1 (2.5) |
| CMV status (Donor/recipient) – no. (%) | | | |
| -/- | 7 (35.0) | 4 (20.0) | 11 (27.5) |
| +/- | 4 (20.0) | 1 (5.0) | 5 (12.5) |
| -/+ | 2 (10.0) | 7 (35.0) | 9 (22.5) |
| +/+ | 7 (35.0) | 8 (40.0) | 15 (37.5) |
| Missing/unknown | 0 | 0 | 0 |

**Supplementary Table 3**

| Differences in inverse Simpson diversity between GVHD prophylaxis arms by timepoint | |
|---|---|
| **Timepoint** | **Ratio of mean inverse Simpson diversity index Tac/MTX:PT-Cy** |
| Baseline (pre-conditioning) | 1.06 |
| Day 0 (pre-infusion) | 1.22 |
| Day 7 | 2.26 |
| Day 14 | 5.42 |
| Day 28 | 3.46 |
| Day 63 | 2.60 |
| Day 98 | 2.18 |
| Day 180 | 3.78 |
| Day 270 | 4.00 |
| Day 365 | 3.57 |
| Day 730 | 4.25 |

**Supplementary Table 4**

| Day 98 landmark analysis of outcomes stratified by TCR diversity | | | | |
|---|---|---|---|---|
| | Estimate at 1 year, % (95% confidence intervals) | | | |
| **Endpoint** | **Higher diversity at day 98** | **Lower diversity at day 98** | **P value** | **Subdistribution Hazard Ratio (95% confidence intervals)** |
| Cumulative incidence of chronic GVHD requiring immunosuppression | 22.6 (14.8—31.4) | 24.9 (16.9—33.7) | 0.73 | 1.11 (0.63—1.96) |
| Cumulative incidence of Grade 2 or 3 Infection | 11.9 (5.8—20.4) | 9.0 (3.9—16.7) | 0.51 | 0.72 (0.27—1.90) |
| Overall survival | 79.2 (71.9—87.4) | 87.0 (80.9—93.6) | 0.15 | 0.61 (0.31—1.20) |
| Cumulative incidence of non-relapse mortality | 9.9 (5.0—16.7) | 5.7 (2.3—11.3) | 0.28 | 0.57 (0.21—1.58) |
| Cumulative incidence of relapse | 18.9 (11.9—27.2) | 9.5 (4.9—16.1) | 0.06 | 0.49 (0.23—1.05) |

**Supplementary Table 5**

| Day 180 landmark analysis of outcomes stratified by TCR diversity | | | | |
|---|---|---|---|---|
| | Estimate at 1 year, % (95% confidence intervals) | | | |
| **Endpoint** | **Higher diversity at day 180** | **Lower diversity at day 180** | **P value** | **Subdistribution Hazard Ratio (95% confidence intervals)** |
| Cumulative incidence of chronic GVHD requiring immunosuppression | 18.3 (10.8—27.3) | 16.7 (9.4—25.9) | 0.73 | 0.88 (0.42—1.84) |
| Cumulative incidence of Grade 2 or 3 Infection | 8.6 (3.5—16.6) | 4.6 (1.2—11.7) | 0.31 | 0.49 (0.12—1.94) |
| Overall survival | 90.5 (84.8—96.6) | 95.9 (92.1—99.9) | 0.16 | 0.43 (0.13—1.41) |
| Cumulative incidence of non-relapse mortality | 2.2 (0.4—7.0) | 3.2 (0.9—8.3) | 0.69 | 1.44 (0.24—8.56) |
| Cumulative incidence of relapse | 13.4 (7.3—21.3) | 5.4 (2.0—11.4) | 0.07 | 0.39 (0.14—1.10) |

**Supplementary Table 6**

| Proportions of GVHD prophylaxis regimens in each TCR diversity group | | |
|---|---|---|
| **Treatment arm – no. (%)** | **Higher diversity at day 14 (N=140)** | **Lower diversity at day 14 (N=141)** |
| **PT-Cy** | 51 (36) | 93 (66) |
| **Tac/MTX** | 89 (64) | 48 (34) |

**Supplementary Table 7**

| Potential covariates for TCR diversity by GVHD prophylaxis arm model | |
|---|---|
| **Potential covariate** | **P value for stepwise variable selection** |
| Gender | 0.18 |
| Race | 0.80 |
| Age | 0.32 |
| Primary disease | 0.23 |
| Conditioning regimen | 0.11 |
| Disease Risk Index | 0.71 |
| Degree of HLA matching | 0.96 |

### Supplementary Figure 1

**Gating strategy**

**Panel #1**

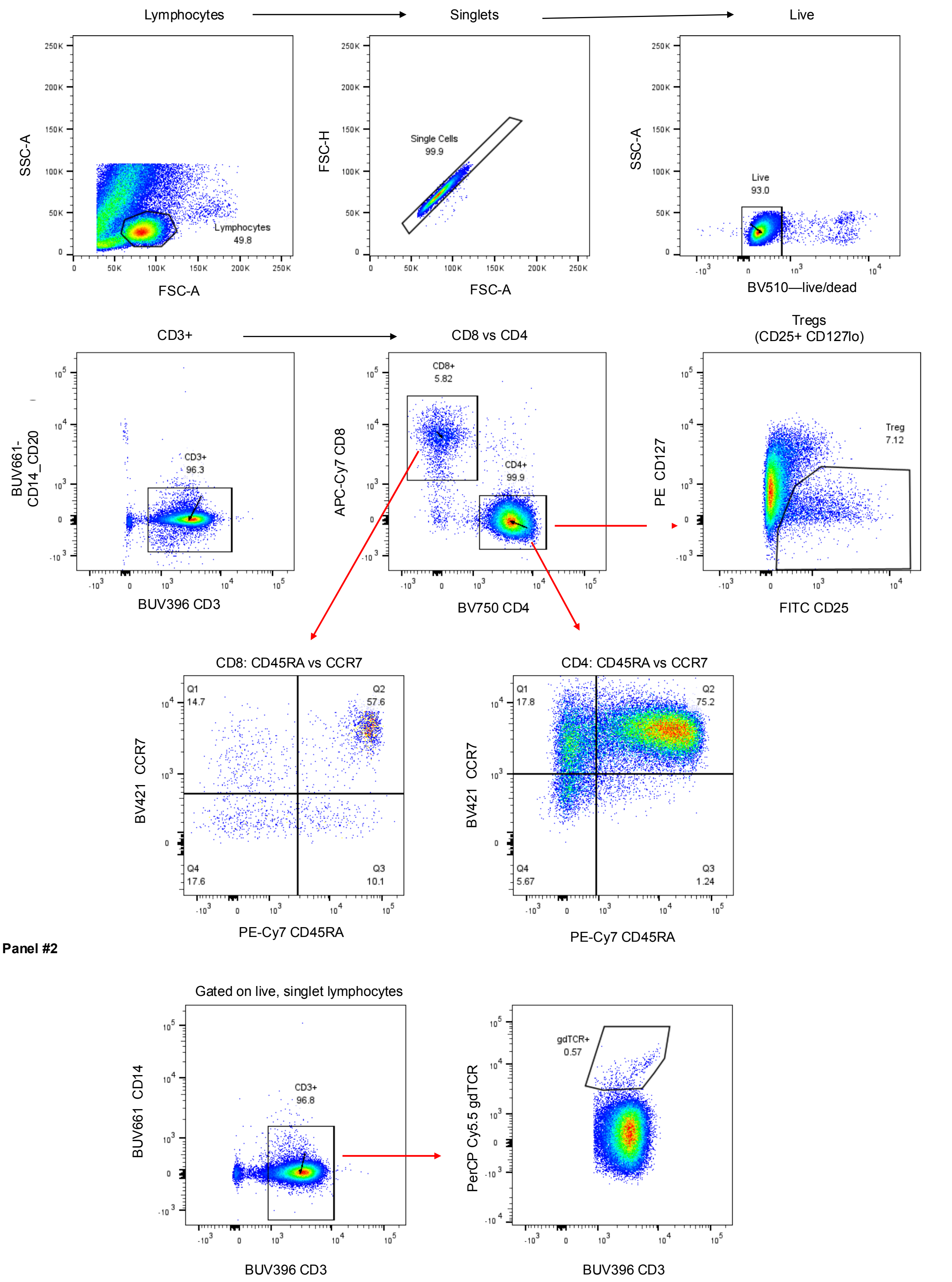


Patient 311 (PT-Cy), Day 7
